# Anemia is a risk factor for rehospitalization after SARS-CoV-2 clearance

**DOI:** 10.1101/2020.12.02.20242958

**Authors:** Patrick Lenehan, Eshwan Ramudu, AJ Venkatakrishnan, Gabriela Berner, Reid McMurry, John C. O’Horo, Andrew D. Badley, William Morice, John Halamka, Venky Soundararajan

**Author notes:** These authors contributed equally (Patrick Lenehan, Eshwan Ramudu).

## Abstract

**Background:** As the number of new and recovering COVID-19 cases continues to rise, it has become evident that patients can experience symptoms and complications after viral clearance. Clinical biomarkers characterizing patients who are likely to experience these prolonged effects are unknown.

**Methods:** We conducted a retrospective study to compare longitudinal lab test measurements (hemoglobin, hematocrit, estimated glomerular filtration rate, serum creatinine, and blood urea nitrogen) in patients rehospitalized after PCR-confirmed SARS-CoV-2 clearance (n=104) versus patients not rehospitalized after viral clearance (n=278).

**Findings:** Compared to patients who were not rehospitalized after PCR-confirmed viral clearance, those who were rehospitalized had lower median hemoglobin levels in the year prior to COVID-19 diagnosis (cohen’s D = -0.50; p=1.2×10^−3^) and during the active infection window (cohen’s D = -0.71; p=4.6×10^−8^). Patients hospitalized after viral clearance were also more likely to be diagnosed with moderate or severe anemia during the active infection window (OR = 2.18; p = 4.99×10^−9^).

**Conclusions:** The occurrence of moderate or severe anemia in hospitalized COVID-19 patients is strongly associated with rehospitalization after viral clearance. Whether interventions to mitigate anemia can improve long term outcomes of COVID-19 patients should be further investigated.

**Funding:** This study was funded by nference.

## Introduction

Since the first diagnosed case of COVID-19 in December 2019, over 95 million people have been infected with SARS-CoV-2 worldwide resulting in over 2 million deaths. While significant progress has been made in understanding the pathogenesis of COVID-19, including the rapid development and clinical rollout of multiple vaccines (Folegatti *et al*., 2020, Jackson *et al*., 2020, Corbett *et al*., 2020, Mercado *et al*., 2020, Bos *et al*., 2020, Mulligan *et al*., 2020) along with detailed characterizations of the SARS-CoV-2 entry receptor ACE2 (Venkatakrishnan *et al*., 2020, Anand *et al*., 2020, Ziegler *et al*., 2020, Zhao *et al*., 2020, Singh *et al*., 2020), there are still few options available for effective treatment of patients with severe COVID-19. Further, as the pandemic has progressed, there have been reports of long-lasting effects of COVID-19 even in patients who did not experience a severe disease course during their active period of infection (Yelin *et al*., 2020, del Rio *et al*., 2020, Carfì *et al*., 2020). However, the clinical, molecular, and demographic biomarkers characterizing patients who are more likely to experience these lasting effects after clearing SARS-CoV-2 are not yet known.

The need to answer such questions during the rapidly evolving COVID-19 pandemic has emphasized the requirement for tools facilitating real-time analysis of patient data as it is obtained and stored in large electronic health records (EHR) systems. Specifically, clinical research efforts to understand the features defining COVID-19 patients, or subsets thereof, fundamentally require reliable systems that enable (1) conversion of unstructured information (e.g., patients notes written by healthcare professionals) into structured formats suitable for downstream analysis and (2) temporal alignment and integration of such unstructured data with the already structured information available in EHR databases (e.g., lab test results, disease diagnosis codes).

With these requirements in mind, we have previously reported the development of augmented curation methods that enable the rapid creation and comparison of defined cohorts of COVID-19 patients within a large EHR system (Pawlowski *et al*., 2020, Wagner *et al*., 2020). Here we expand on our prior textual sentiment-based analysis (Pawlowski *et al*., 2020) to understand the clinical features of patients likely to experience lasting effects of COVID-19, and we find that lab tests support our previously derived hypothesis that anemia and kidney malfunction during active SARS-CoV-2 infection may serve as biomarkers of patients who are more likely to be rehospitalized after PCR-confirmed viral clearance.

## Results

### Longitudinal analysis of laboratory measurements provides a framework to test hypotheses derived from unstructured electronic health records

Using NLP-based extraction of phenotypes from a large EHR system, we previously reported that COVID-19 patients who are rehospitalized after PCR-confirmed viral clearance were more likely to experience anemia and acute kidney injury (AKI) in the year prior to their diagnosis and during their PCR-positive phase of COVID-19 compared to patients who were not rehospitalized after clearance of SARS-CoV-2 (Pawlowski *et al*., 2020). Here, we sought to evaluate whether diagnostic lab tests for AKI and anemia corroborate these phenotypic associations in a larger patient cohort.

We split the set of hospitalized COVID-19 patients with confirmed viral clearance (n=382) into two groups: (1) post-clearance hospitalized (“PCH”; n=104) and (2) post-clearance non-hospitalized (“PCNH”; n=278), where viral clearance was defined by two consecutive negative SARS-CoV-2 PCR tests following a positive test (**Figure 1A-B**). A demographic summarization of these two cohorts is provided in **Table 1**. We then compared a set of selected lab test results between these cohorts during two time windows: (1) the year prior to COVID-19 diagnosis (“pre-COVID phase”) and (2) the time during which each patient was SARS-CoV-2 positive according to their PCR results (“SARS-CoV-2^+^ phase”). Given our previous EHR-based findings, we considered both anemia-related and kidney function lab tests including hemoglobin, hematocrit, estimated glomerular filtration rate (eGFR), serum creatinine, and serum blood urea nitrogen (BUN) levels (**Figure 1C**).

**Table 1.** Demographics and clinical characteristics of study cohorts, including patients who were and who were not rehospitalized after PCR-confirmed clearance of SARS-CoV-2. Each demographic variable or clinical characteristic was tested for difference in proportion with a Fisher Exact test or a difference in magnitude (for continuous variables) using a Mann-Whitney U test, and p-values were corrected for multiple testing using the Benjamini-Hochberg (BH) correction.

| Demographic or Clinical Characteristic | Hospitalized Post-Clearance<br>("PCH Cohort") | Non-Hospitalized Post-Clearance<br>("PCNH Cohort") | BH-adjusted p-value |
| --- | --- | --- | --- |
| Total number of patients, *N* | 104 | 278 |  |
| Age, years (*standard deviation*) | 59.2 (18.4) | 59.3 (17.5) | 1.000 |
| Sex |  |  |  |
| - Female, *n (%)* | 50 (48.1%) | 118 (42.4%) | 0.639 |
| - Male, *n (%)* | 54 (51.9%) | 160 (57.6%) | 0.639 |
| Race |  |  |  |
| - White, *n (%)* | 78 (75.0%) | 184 (66.2%) | 0.433 |
| - Asian, *n (%)* | 5 (4.8%) | 22 (7.9%) | 0.639 |
| - Black, *n (%)* | 6 (5.8%) | 38 (13.7%) | 0.187 |
| - Other, *n (%)* | 15 (14.4%) | 34 (12.2%) | 0.728 |
| Ethnicity |  |  |  |
| - Hispanic, *n (%)* | 16 (15.4%) | 53 (19.1%) | 0.686 |
| - Non-Hispanic, *n (%)* | 85 (81.7%) | 218 (78.4%) | 0.728 |
| - Other, *n (%)* | 3 (2.9%) | 7 (2.5%) | 1.000 |
| Relative Cleared Date, days (*standard deviation*) | 39.5 (39.4) | 32.4 (29.1) | 0.639 |
| ICU Admission, *n (%)* | 48 (46%) | 73 (26%) | 0.004 |

**Figure 1.**
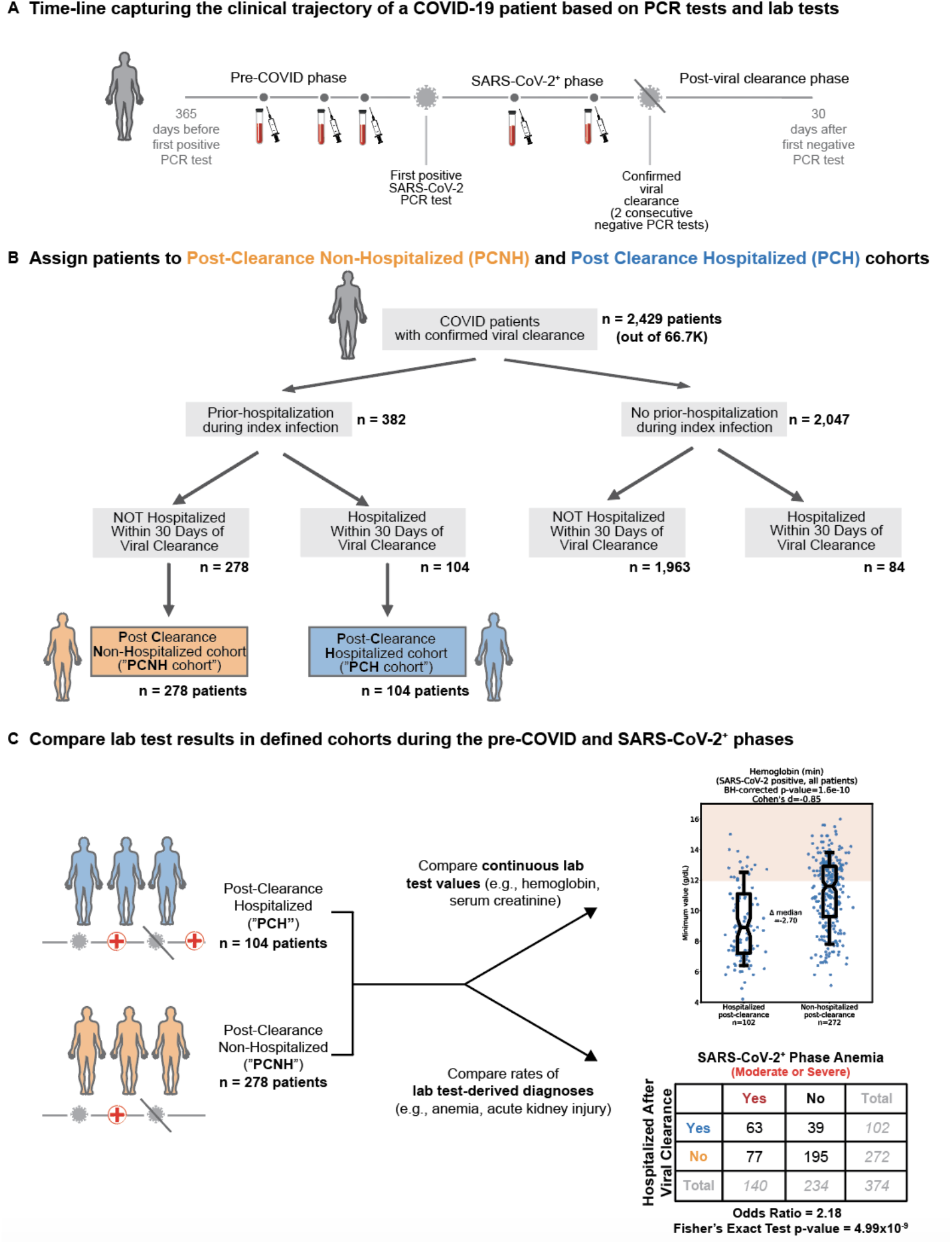
Schematic summarizing cohort creation and lab test analyses. (A) Time intervals (“phases”) were defined relative to SARS-CoV-2 PCR testing results. (B) Of the 2,429 patients who were diagnosed by PCR with COVID-19 and subsequently confirmed to have cleared SARS-CoV-2 with two consecutive negative tests, we created two cohorts: (1) patients who were hospitalized during their index infection and not hospitalized after confirmed viral clearance (post-clearance non-hospitalized, or “PCNH”; n=278), and (2) patients who were hospitalized during their index infection and rehospitalized within 90 days of confirmed viral clearance (post clearance hospitalized, or “PCH”; n=104). (C) A defined set of anemia and kidney related lab test measurements were compared between the PCH and PCNH cohorts in the pre-COVID and SARS-CoV-2^+^ intervals.

### Rehospitalized patients show pathologic alterations in hemoglobin, hematocrit, eGFR, and BUN before COVID-19 diagnosis and during active infection

For each patient, we first considered the median values of each lab test over the designated interval. Consistent with our previous augmented curation derived findings, PCH patients had significantly lower median hemoglobin and hematocrit levels in both the pre-COVID phase (cohen’s D = -0.50, p=1.2×10^−3^; cohen’s D = -0.48, p=2.5×10^−3^) and the SARS-CoV-2^+^ phase (cohen’s D = -0.71, p=4.6×10^−8^; cohen’s D = -0.69, p=8.5×10^−8^) (**Table 2, Figures 2A-D**). Further, PCH patients had lower median eGFR and higher median BUN levels during the pre-COVID phase (cohen’s D = -0.46, p = 0.02; cohen’s D = -0.45, p=1.2×10^−3^) and the SARS-CoV-2^+^ phase (cohen’s D = 0.46, p = 0.01; cohen’s D = 0.42, p=8.9×10^−6^) (**Table 2, Figure S1**).

**Table 2.** Analysis of median values for all selected lab tests in pre-COVID and SARS-CoV-2^+^ phases, including both male and female patients. Entries are sorted in order of statistical significance by the BH-adjusted Mann Whitney U-test p-value. Abbreviations are defined as follows: Hgb - hemoglobin; Hct - hematocrit; eGFR - estimated glomerular filtration rate; Cr - creatinine, BUN - blood urea nitrogen; g/DL - grams per deciliter; mL/min/BSA - milliliters per minute normalized for body surface area.

| Test | Units | Phase | N<br>(PCH) | Mean of<br>Medians<br>(PCH) | N<br>(PCNH) | Mean of<br>Medians<br>(PCNH) | Cohen's D | BH-Adj P-val |
| --- | --- | --- | --- | --- | --- | --- | --- | --- |
| Hgb | g/dL | SARS-CoV-2<sup>+</sup> | 102 | 10.70 | 272 | 12.13 | -0.71 | 4.57E-08 |
| Hct | % | SARS-CoV-2<sup>+</sup> | 102 | 32.65 | 272 | 36.61 | -0.69 | 8.46E-08 |
| BUN | mg/dL | SARS-CoV-2<sup>+</sup> | 102 | 26.99 | 272 | 20.85 | 0.42 | 8.87E-06 |
| eGFR | mL/min/BSA | SARS-CoV-2<sup>+</sup> | 83 | 60.55 | 188 | 69.72 | -0.45 | 1.21E-03 |
| Hgb | g/dL | Pre-COVID | 55 | 11.50 | 136 | 12.60 | -0.50 | 1.23E-03 |
| Hct | % | Pre-COVID | 55 | 35.55 | 132 | 38.60 | -0.48 | 2.46E-03 |
| Cr | mg/dL | SARS-CoV-2<sup>+</sup> | 103 | 1.51 | 272 | 1.20 | 0.20 | 5.53E-03 |
| BUN | mg/dL | Pre-COVID | 56 | 25.73 | 133 | 19.80 | 0.46 | 1.33E-02 |
| Cr | mg/dL | Pre-COVID | 57 | 1.95 | 142 | 1.38 | 0.31 | 1.93E-02 |
| eGFR | mL/min/BSA | Pre-COVID | 47 | 56.65 | 109 | 65.92 | -0.46 | 2.45E-02 |

**Figure 2.**
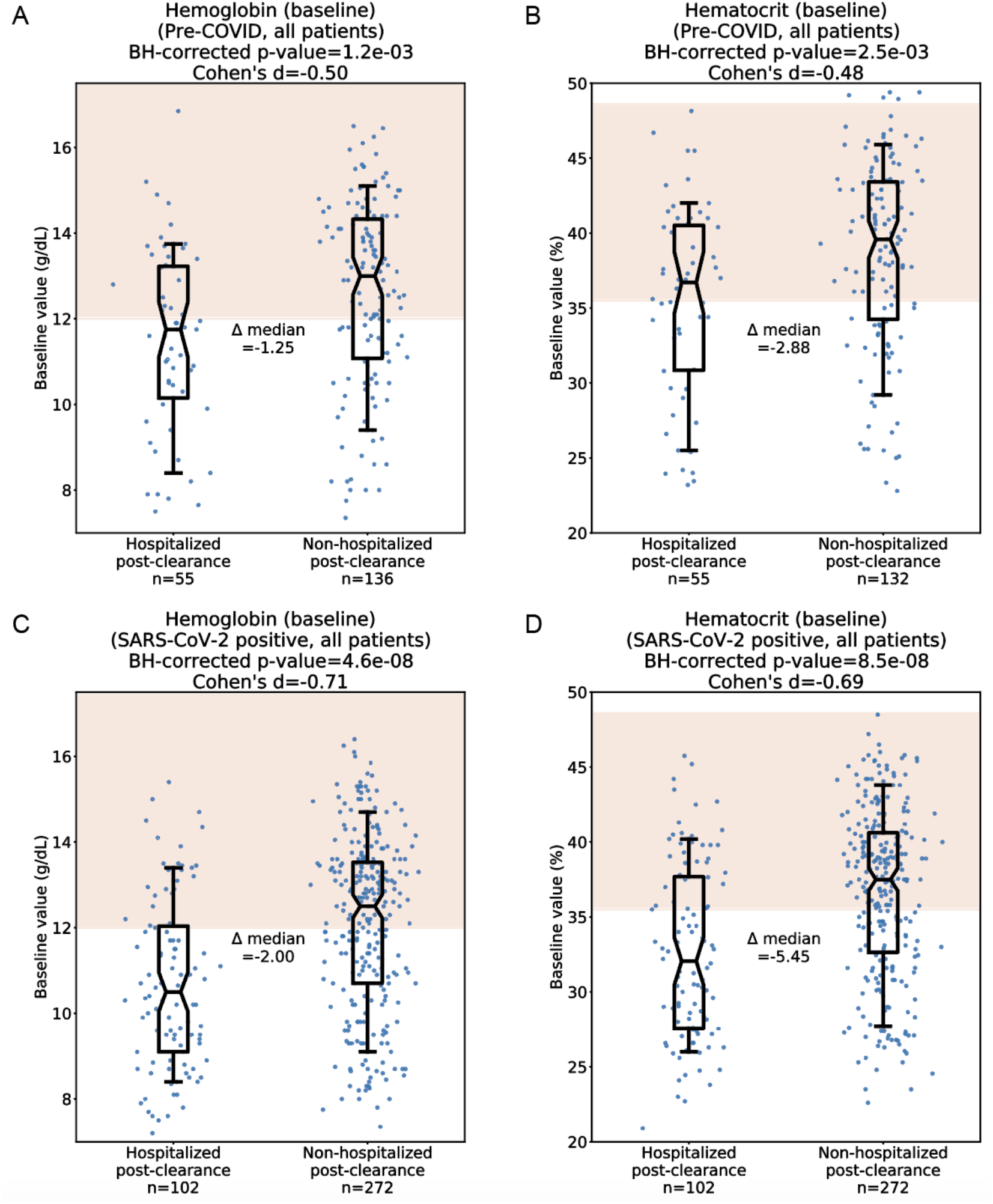
Comparison of median hemoglobin and hematocrit values during the pre-COVID and SARS-CoV-2^+^ phases. (A) Pre-COVID median hemoglobin in the PCH (n=55) and PCNH (n=136) cohorts. (B) Pre-COVID median hematocrit in the PCH (n=55) and PCNH (n=132) cohorts. (C) SARS-CoV-2^+^ median hemoglobin in the PCH (n=102) and PCNH (n=272) cohorts. (D) SARS-CoV-2^+^ median hematocrit in the PCH (n=102) and PCNH (n=272) cohorts. Red shading indicates normal ranges for hemoglobin and hematocrit, spanning from the lower limit of normal for females (12 g/dL hemoglobin, 35.5% hematocrit) to the upper limit of normal for males (17.5 g/dL hemoglobin, 48.6% hematocrit). For each comparison, statistics shown include the number of patients analyzed, Cohen’s D, BH-corrected Mann Whitney U test p-value, and the difference of medians between the two cohorts. Box and whisker plots depict median and IQR along with the 10th and 90th percentiles.

We also tested whether extreme (i.e. minimum or maximum) values of a given lab test over the designated periods varied between PCH and PCNH patients, as a measure of central tendency (e.g., median) may fail to capture a single occurrence of phenotypes such as anemia or AKI. PCH patients had lower minimum values of hemoglobin, hematocrit, and eGFR in both the pre-COVID phase (cohen’s D = -0.49, p=2.8×10^−3^; cohen’s D = -0.45, p=3.0×10^−3^; cohen’s D = - 0.57, p=3.0×10^−3^) and the SARS-CoV-2^+^ phase (cohen’s D = -0.85, p=1.6×10^−10^; cohen’s D = - 0.79, p=1.2×10^−9^; cohen’s D = -0.51, p=4.4×10^−4^). They also had higher maximum serum BUN levels during both the pre-COVID phase (cohen’s D = 0.50, p=6.6×10^−4^) and the SARS-CoV-2^+^ phase (cohen’s D = 0.60, p=5.2×10^−8^) (**Tables 3-4, Figure 3 and Figure S2**).

**Table 3.** Analysis of minimum values for all selected lab tests in pre-COVID and SARS-CoV-2^+^ phases, including both male and female patients. Entries are sorted in order of statistical significance by the BH-adjusted Mann Whitney U-test p-value. Abbreviations are defined as follows: Hgb - hemoglobin; Hct - hematocrit; eGFR - estimated glomerular filtration rate; Cr - creatinine, BUN - blood urea nitrogen; g/DL - grams per deciliter; mL/min/BSA - milliliters per minute normalized for body surface area.

| Test | Units | Phase | N<br>(PCH) | Mean of<br>Minima<br>(PCH) | N<br>(PCNH) | Mean of<br>Minima<br>(PCNH) | Cohen's D | BH-Adj P-val |
| --- | --- | --- | --- | --- | --- | --- | --- | --- |
| Hgb | g/dL | SARS-CoV-2<sup>+</sup> | 102 | 9.18 | 272 | 11.17 | -0.85 | 1.61E-10 |
| Hct | % | SARS-CoV-2<sup>+</sup> | 102 | 28.98 | 272 | 34.03 | -0.79 | 1.22E-09 |
| eGFR | mL/min/BSA | SARS-CoV-2<sup>+</sup> | 83 | 45.70 | 188 | 57.56 | -0.51 | 4.38E-04 |
| Hgb | g/dL | Pre-COVID | 55 | 10.33 | 136 | 11.65 | -0.49 | 2.82E-03 |
| eGFR | mL/min/BSA | Pre-COVID | 47 | 44.72 | 109 | 56.68 | -0.57 | 2.99E-03 |
| Hct | % | Pre-COVID | 55 | 32.26 | 132 | 35.82 | -0.45 | 3.01E-03 |
| BUN | mg/dL | SARS-CoV-2<sup>+</sup> | 102 | 16.01 | 272 | 14.48 | 0.14 | 1.04E-01 |
| Cr | mg/dL | Pre-COVID | 57 | 1.48 | 142 | 1.17 | 0.23 | 1.20E-01 |
| Cr | mg/dL | SARS-CoV-2<sup>+</sup> | 103 | 1.10 | 272 | 0.97 | 0.12 | 2.54E-01 |
| BUN | mg/dL | Pre-COVID | 56 | 16.99 | 133 | 14.62 | 0.27 | 4.23E-01 |

**Table 4.** Analysis of maximum values for all selected lab tests in pre-COVID and SARS-CoV-2^+^ phases, including both male and female patients. Entries are sorted in order of statistical significance by the BH-adjusted Mann Whitney U-test p-value. Abbreviations are defined as follows: Hgb - hemoglobin; Hct - hematocrit; eGFR - estimated glomerular filtration rate; Cr - creatinine, BUN - blood urea nitrogen; g/DL - grams per deciliter; mL/min/BSA - milliliters per minute normalized for body surface area.

| Test | Units | Phase | N (PCH) | Mean of Maxima (PCH) | N (PCNH) | Mean of Maxima (PCNH) | Cohen's D | BH-Adj P-val |
| --- | --- | --- | --- | --- | --- | --- | --- | --- |
| BUN | mg/dL | SARS-CoV-2<sup>+</sup> | 102 | 45.21 | 272 | 30.04 | 0.60 | 5.23E-08 |
| Hct | % | SARS-CoV-2<sup>+</sup> | 102 | 37.50 | 272 | 40.32 | -0.48 | 1.50E-04 |
| Cr | mg/dL | SARS-CoV-2<sup>+</sup> | 103 | 2.22 | 272 | 1.56 | 0.30 | 1.50E-04 |
| BUN | mg/dL | Pre-COVID | 56 | 40.23 | 133 | 27.77 | 0.50 | 6.59E-04 |
| Cr | mg/dL | Pre-COVID | 57 | 2.58 | 142 | 1.67 | 0.37 | 1.04E-03 |
| Hgb | g/dL | SARS-CoV-2<sup>+</sup> | 102 | 13.40 | 272 | 13.75 | -0.10 | 3.85E-03 |
| Hgb | g/dL | Pre-COVID | 55 | 12.97 | 136 | 13.53 | -0.30 | 1.78E-02 |
| Hct | % | Pre-COVID | 55 | 40.05 | 132 | 41.36 | -0.24 | 6.26E-02 |
| eGFR | mL/min/BSA | SARS-CoV-2<sup>+</sup> | 83 | 72.51 | 188 | 80.14 | -0.41 | 9.20E-02 |
| eGFR | mL/min/BSA | Pre-COVID | 47 | 67.13 | 109 | 74.68 | -0.38 | 2.42E-01 |

**Figure 3.**
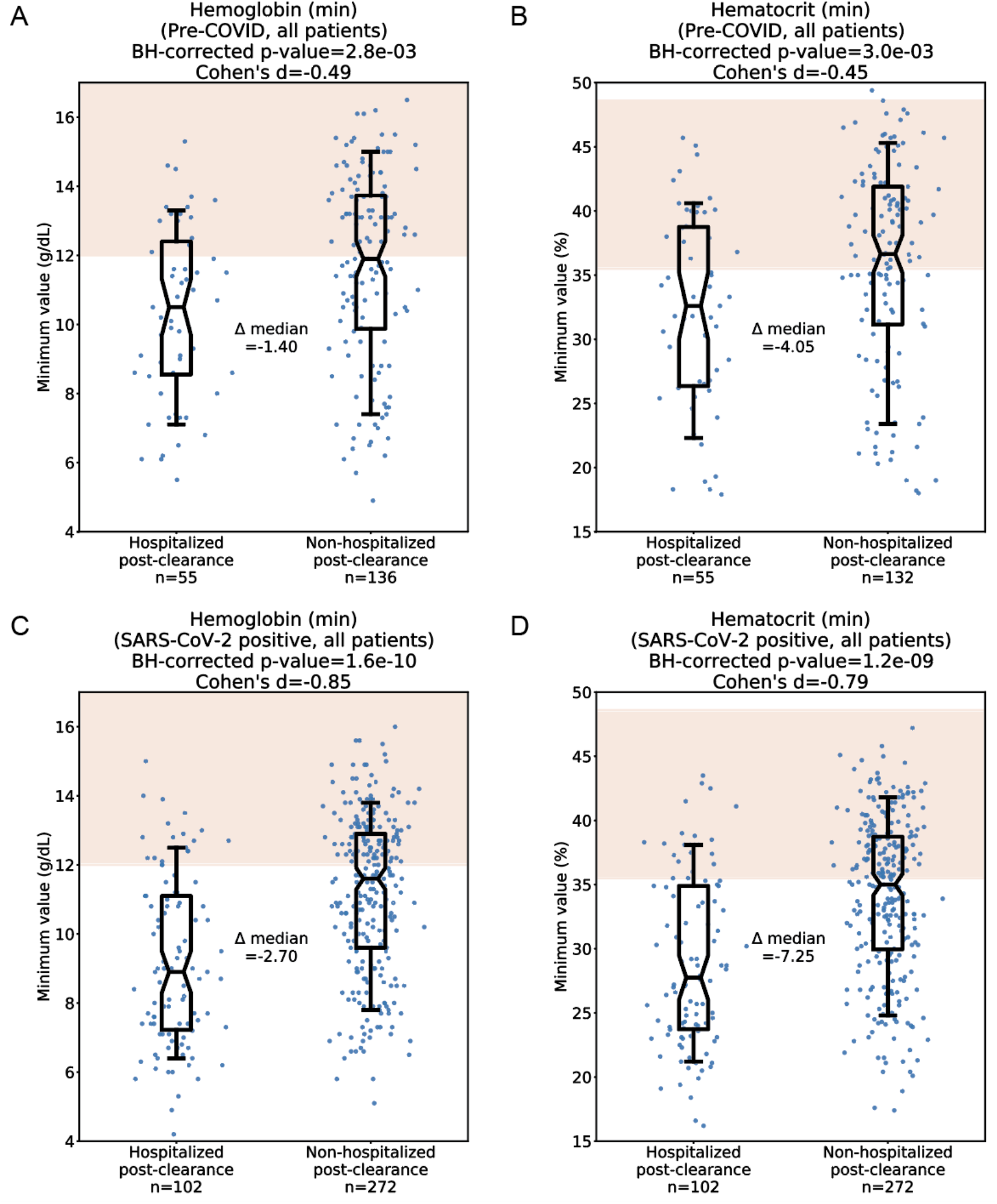
Comparison of minimum values for hemoglobin and hematocrit in the pre-COVID and SARS-CoV-2^+^ intervals. (A) Pre-COVID minimum hemoglobin in the PCH (n=55) and PCNH (n=136) cohorts. (B) Pre-COVID minimum hematocrit in the PCH (n=55) and PCNH (n=132) cohorts. (C) SARS-CoV-2^+^ minimum hemoglobin in the PCH (n=102) and PCNH (n=272) cohorts.(D) SARS-CoV-2^+^ minimum hematocrit in the PCH (n=102) and PCNH (n=272) cohorts. Red shading indicates normal ranges for hemoglobin and hematocrit as described in Figure 1. For each comparison, statistics shown include the number of patients analyzed, Cohen’s D, BH-corrected Mann Whitney U test p-value, and the difference of medians between the two cohorts. Box and whisker plots depict median and IQR along with the 10th and 90th percentiles.

Taken together, these analyses corroborate our prior textual sentiment-based EHR findings, suggesting that patients who are rehospitalized after SARS-CoV-2 clearance are more likely to have pathologically altered anemia-related and renal function lab tests both prior to and during SARS-CoV-2 infection.

### Post-clearance rehospitalized patients have lower hemoglobin and hematocrit before and during SARS-CoV-2 infection regardless of sex

As males and females have different normal ranges of hemoglobin and hematocrit, we performed sex-split subanalyses of anemia-related lab tests similar to those described above. Patient-level median hemoglobin and hematocrit during the pre-COVID phase were significantly lower in both the female (cohen’s D = -0.66, p=0.01; cohen’s D = -0.67, p=0.01) and male (cohen’s D = -0.42, p=0.02; cohen’s D = -0.37, p=0.05) PCH cohorts versus their PCNH counterparts (**Tables 5-6, Figures 4A-D**). These trends were even stronger during the SARS-CoV-2^+^ phase among both the female (cohen’s D = -0.85; p= 7.0×10^−6^; cohen’s D = -0.91, p=7.0×10^−6^) and male (cohen’s D = -0.60; p=8.2×10^−4^; cohen’s D = -0.53, p=1.8×10^−3^) cohorts (**Tables 5-6, Figures 4E-H**).

**Table 5.**
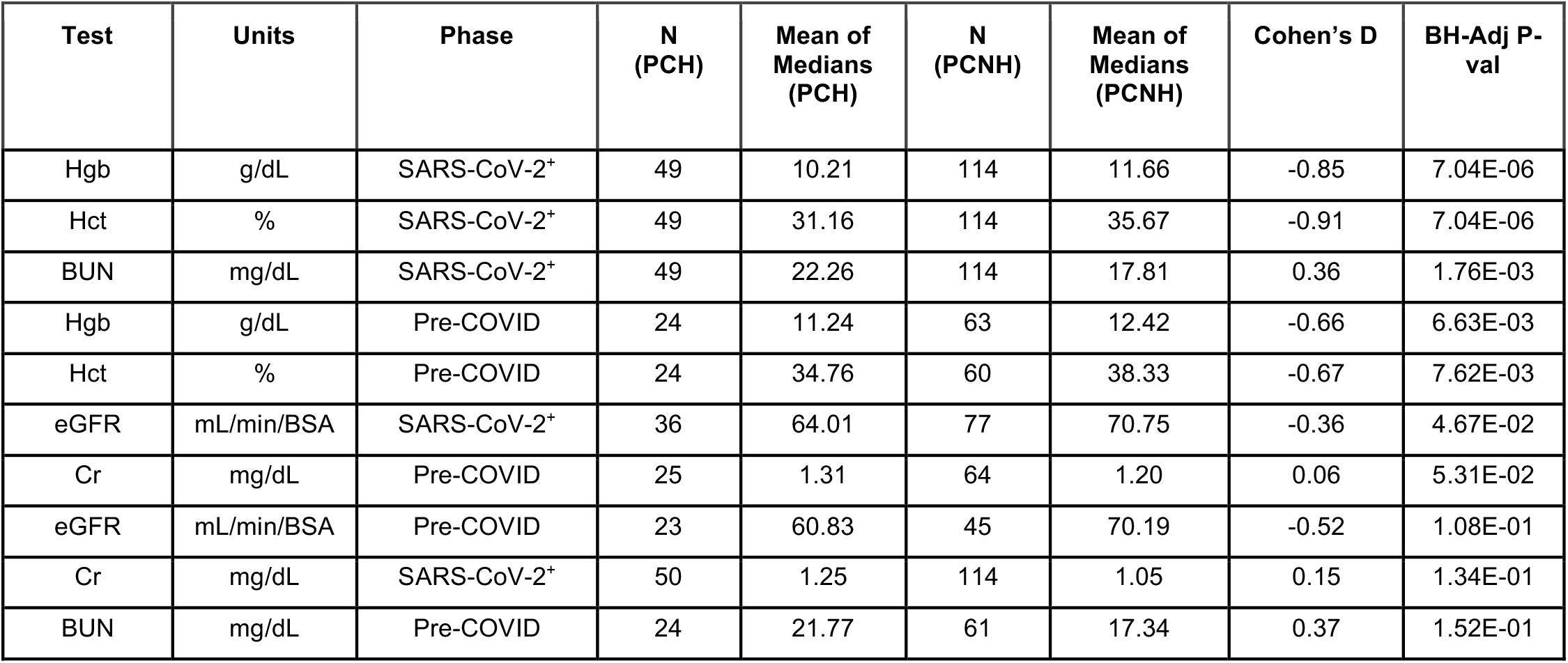
Sex-split analysis of median values for all selected lab tests in female patients during the pre-COVID and SARS-CoV-2^+^ phases. Entries are sorted in order of statistical significance by the BH-adjusted Mann Whitney U-test p-value. Abbreviations are defined as follows: Hgb - hemoglobin; Hct - hematocrit; eGFR - estimated glomerular filtration rate; Cr - creatinine, BUN - blood urea nitrogen; g/DL - grams per deciliter; mL/min/BSA - milliliters per minute normalized for body surface area.

**Table 6.** Sex-split analysis of median values for all selected lab tests in male patients during the pre-COVID and SARS-CoV-2^+^ phases. Entries are sorted in order of statistical significance by the BH-adjusted Mann Whitney U-test p-value. Abbreviations are defined as follows: Hgb - hemoglobin; Hct - hematocrit; eGFR - estimated glomerular filtration rate; Cr - creatinine, BUN - blood urea nitrogen; g/DL - grams per deciliter; mL/min/BSA - milliliters per minute normalized for body surface area.

| Test | Units | Phase | N (PCH) | Mean of Medians (PCH) | N (PCNH) | Mean of Medians (PCNH) | Cohen's D | BH-Adj P-val |
| --- | --- | --- | --- | --- | --- | --- | --- | --- |
| BUN | mg/dL | SARS-CoV-2<sup>+</sup> | 53 | 31.36 | 158 | 23.05 | 0.55 | 5.40E-04 |
| Hgb | g/dL | SARS-CoV-2<sup>+</sup> | 53 | 11.15 | 158 | 12.47 | -0.60 | 8.20E-04 |
| Cr | mg/dL | SARS-CoV-2<sup>+</sup> | 53 | 1.75 | 158 | 1.30 | 0.28 | 1.75E-03 |
| Hct | % | SARS-CoV-2<sup>+</sup> | 53 | 34.03 | 158 | 37.29 | -0.53 | 1.79E-03 |
| eGFR | mL/min/BSA | SARS-CoV-2<sup>+</sup> | 47 | 57.89 | 111 | 69.00 | -0.53 | 6.05E-03 |
| Hgb | g/dL | Pre-COVID | 31 | 11.70 | 73 | 12.75 | -0.42 | 1.88E-02 |
| BUN | mg/dL | Pre-COVID | 32 | 28.70 | 72 | 21.89 | 0.52 | 1.88E-02 |
| Hct | % | Pre-COVID | 31 | 36.16 | 72 | 38.83 | -0.37 | 5.19E-02 |
| eGFR | mL/min/BSA | Pre-COVID | 24 | 52.65 | 64 | 62.91 | -0.49 | 6.35E-02 |
| Cr | mg/dL | Pre-COVID | 32 | 2.44 | 78 | 1.52 | 0.48 | 7.43E-02 |

**Figure 4.**
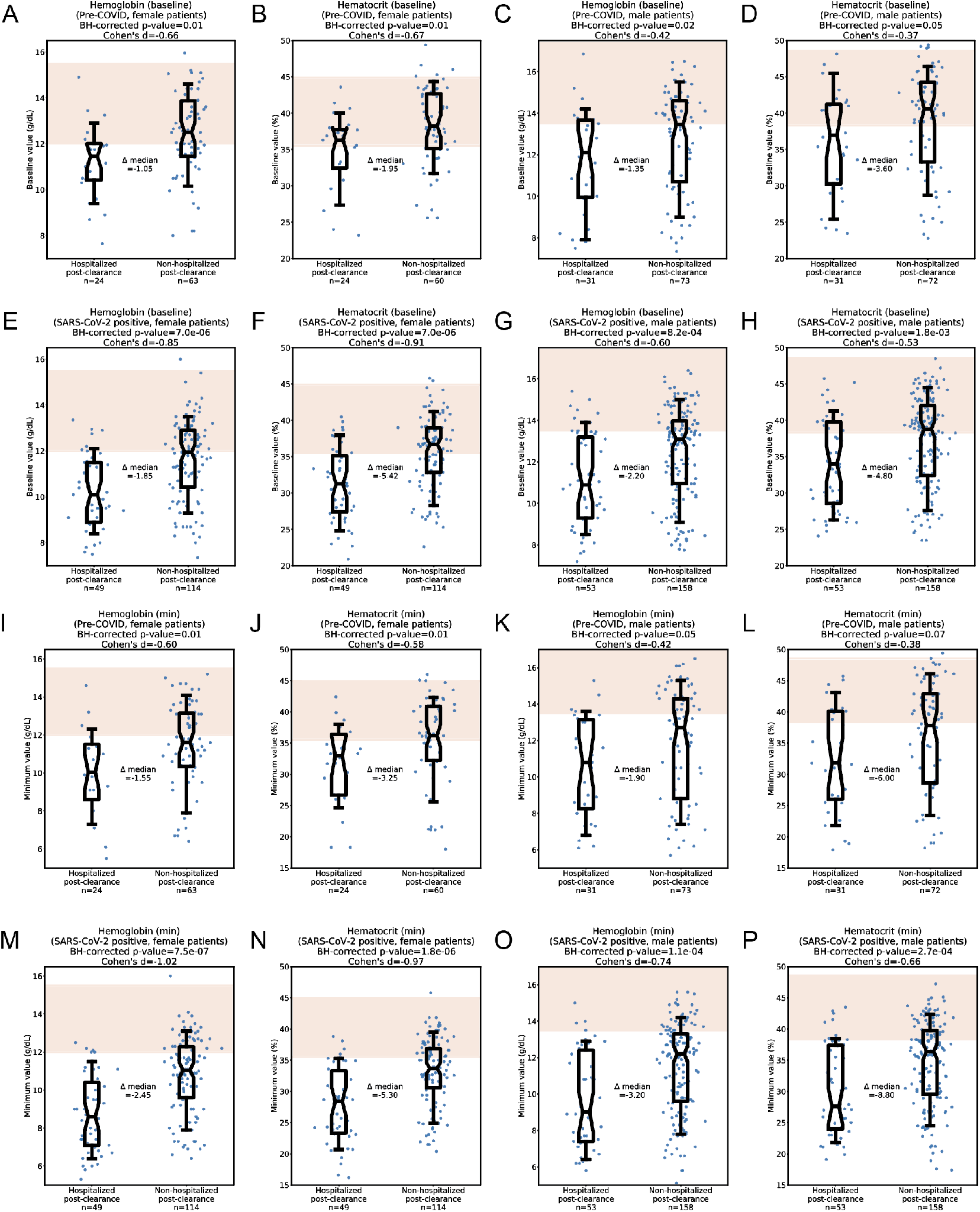
Sex-split analysis of anemia-related lab tests in the pre-COVID and SARS-CoV-2^+^ phases. (A-H) Median values of hemoglobin and hematocrit in the pre-COVID phase (A-D) and SARS-CoV-2^+^ phase (E-H), split to show female patients (A-B, E-F) and male patients (C-D, G-H) separately. (I-P) Minimum values of hemoglobin and hematocrit in the pre-COVID phase (I-L) and SARS-CoV-2^+^ phase (M-P), split to show female patients (I-J, M-N) and male patients (K-L, O-P) separately. Red shading indicates normal ranges for hemoglobin and hematocrit depending on sex (females: 12.0-15.5 g/dL, 35.5-44.9%; males: 12.0-15.5 g/dL, 38.3-48.6%). For each comparison, statistics shown include the number of patients analyzed, Cohen’s D, BH-corrected Mann Whitney U test p-value, and the difference of medians between the two cohorts. Box and whisker plots depict median and IQR along with the 10th and 90th percentiles.

Similarly, in our analysis of extreme values, minimum hemoglobin and hematocrit measurements during the pre-COVID phase tended to be lower in both female (cohen’s D = - 0.60; p=0.01; cohen’s D = -0.58, p=0.01) and male (cohen’s D = -0.42; p=0.05; cohen’s D = -0.38, p=0.07) PCH patients (**Tables 7-8, Figures 4I-L**). These trends were again even stronger in the SARS-CoV-2^+^ phase among both females (cohen’s D = -1.02; p=7.5×10^−7^; cohen’s D = -0.97, p=1.8×10^−6^) and males (cohen’s D = -0.74; p=1.1×10^−4^; cohen’s D = -0.66, p=2.7×10^−4^) (**Tables 7-8, Figures 4M-P**).

**Table 7.**
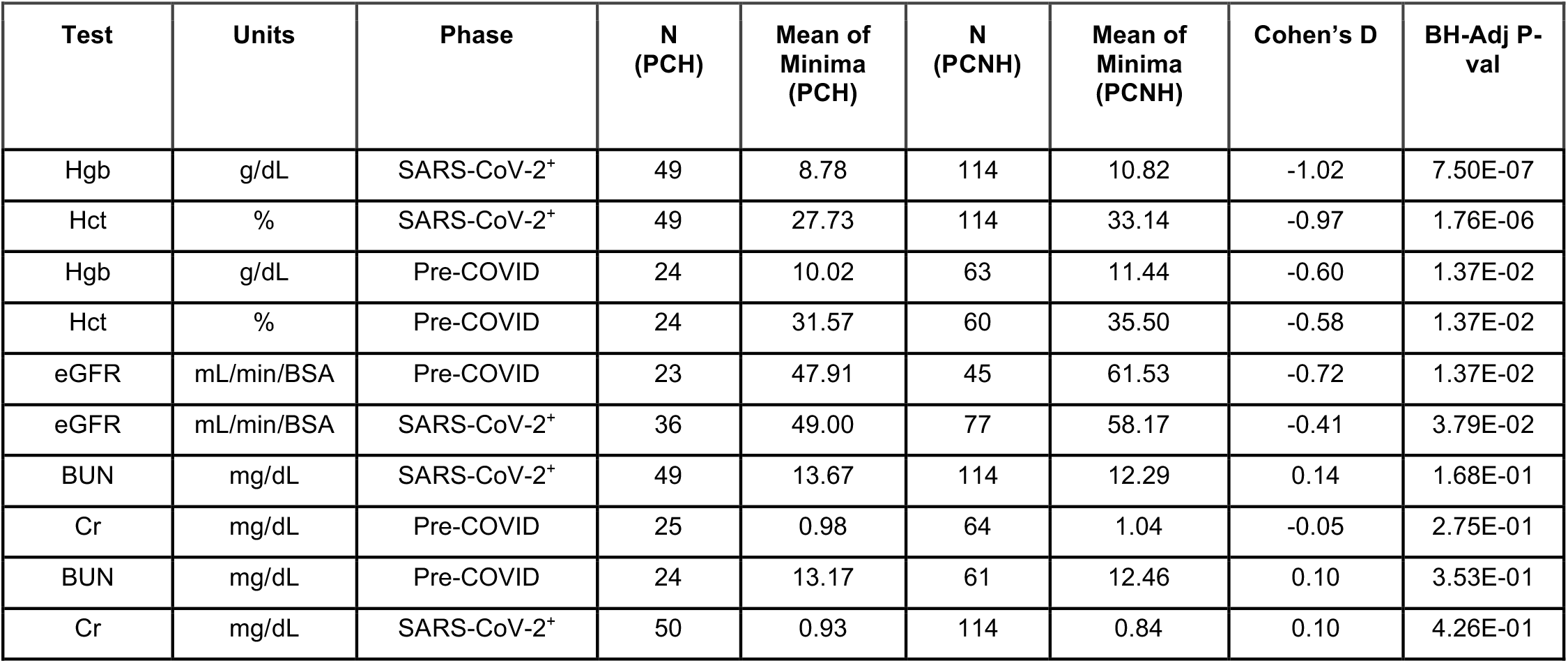
Sex-split analysis of minimum values for all selected lab tests in female patients during the pre-COVID and SARS-CoV-2^+^ phases. Entries are sorted in order of statistical significance by the BH-adjusted Mann Whitney U-test p-value. Abbreviations are defined as follows: Hgb - hemoglobin; Hct - hematocrit; eGFR - estimated glomerular filtration rate; Cr - creatinine, BUN - blood urea nitrogen; g/DL - grams per deciliter; mL/min/BSA - milliliters per minute normalized for body surface area.

| Test | Units | Phase | N<br>(PCH) | Mean of<br>Minima<br>(PCH) | N<br>(PCNH) | Mean of<br>Minima<br>(PCNH) | Cohen's D | BH-Adj P-val |
| --- | --- | --- | --- | --- | --- | --- | --- | --- |
| Hgb | g/dL | SARS-CoV-2<sup>+</sup> | 49 | 8.78 | 114 | 10.82 | -1.02 | 7.50E-07 |
| Hct | % | SARS-CoV-2<sup>+</sup> | 49 | 27.73 | 114 | 33.14 | -0.97 | 1.76E-06 |
| Hgb | g/dL | Pre-COVID | 24 | 10.02 | 63 | 11.44 | -0.60 | 1.37E-02 |
| Hct | % | Pre-COVID | 24 | 31.57 | 60 | 35.50 | -0.58 | 1.37E-02 |
| eGFR | mL/min/BSA | Pre-COVID | 23 | 47.91 | 45 | 61.53 | -0.72 | 1.37E-02 |
| eGFR | mL/min/BSA | SARS-CoV-2<sup>+</sup> | 36 | 49.00 | 77 | 58.17 | -0.41 | 3.79E-02 |
| BUN | mg/dL | SARS-CoV-2<sup>+</sup> | 49 | 13.67 | 114 | 12.29 | 0.14 | 1.68E-01 |
| Cr | mg/dL | Pre-COVID | 25 | 0.98 | 64 | 1.04 | -0.05 | 2.75E-01 |
| BUN | mg/dL | Pre-COVID | 24 | 13.17 | 61 | 12.46 | 0.10 | 3.53E-01 |
| Cr | mg/dL | SARS-CoV-2<sup>+</sup> | 50 | 0.93 | 114 | 0.84 | 0.10 | 4.26E-01 |

**Table 8.**
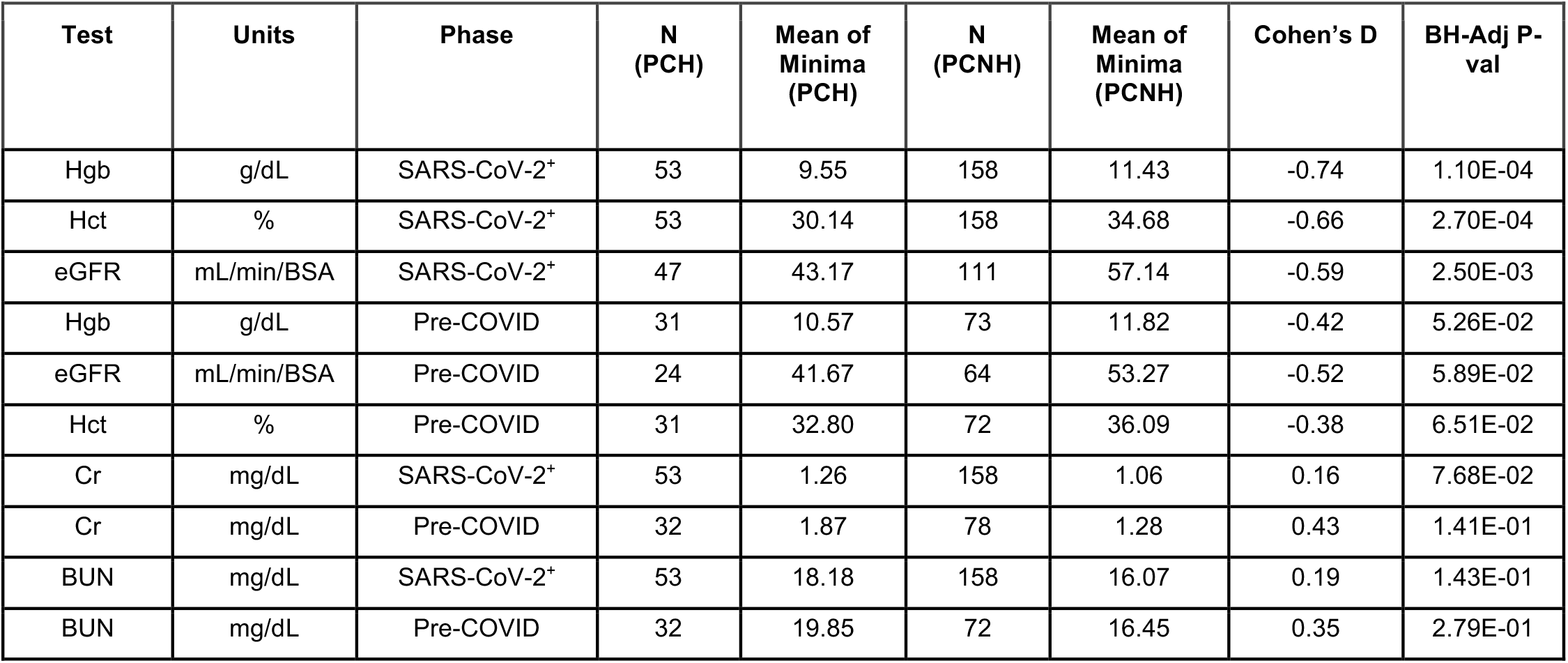
Sex-split analysis of minimum values for all selected lab tests in male patients during the pre-COVID and SARS-CoV-2^+^ phases. Entries are sorted in order of statistical significance by the BH-adjusted Mann Whitney U-test p-value. Abbreviations are defined as follows: Hgb - hemoglobin; Hct - hematocrit; eGFR - estimated glomerular filtration rate; Cr - creatinine, BUN - blood urea nitrogen; g/DL - grams per deciliter; mL/min/BSA - milliliters per minute normalized for body surface area.

### Post-clearance rehospitalized patients are more likely to have experienced anemia and AKI before COVID-19 diagnosis and during active infection

We next evaluated whether outright anemia occurred more frequently in the PCH cohort than the PCNH cohort (see **Methods** and **Figure 5A**). Anemia was indeed observed more frequently in the PCH cohort during both the pre-COVID phase (39/55 [71%] vs. 61/136 [45%]; OR=1.58; p=0.001) and the SARS-CoV-2^+^ phase (93/102 [91%] vs. 202/272 [74%]; OR=1.23; p=2.02×10^−4^) (**Figures 5B-C**). Further, the prevalence of moderate or severe anemia was higher in the PCH cohorts during both the pre-COVID phase (13/55 [31%] vs. 19/136 [14%]; OR=1.69; p=0.13) and the SARS-CoV-2^+^ phase (63/102 [62%] vs. 77/272 [28%]; OR=2.182; p=4.99×10^−9^), although the former observation did not reach statistical significance (**Figures 5D-E**).

**Figure 5.**
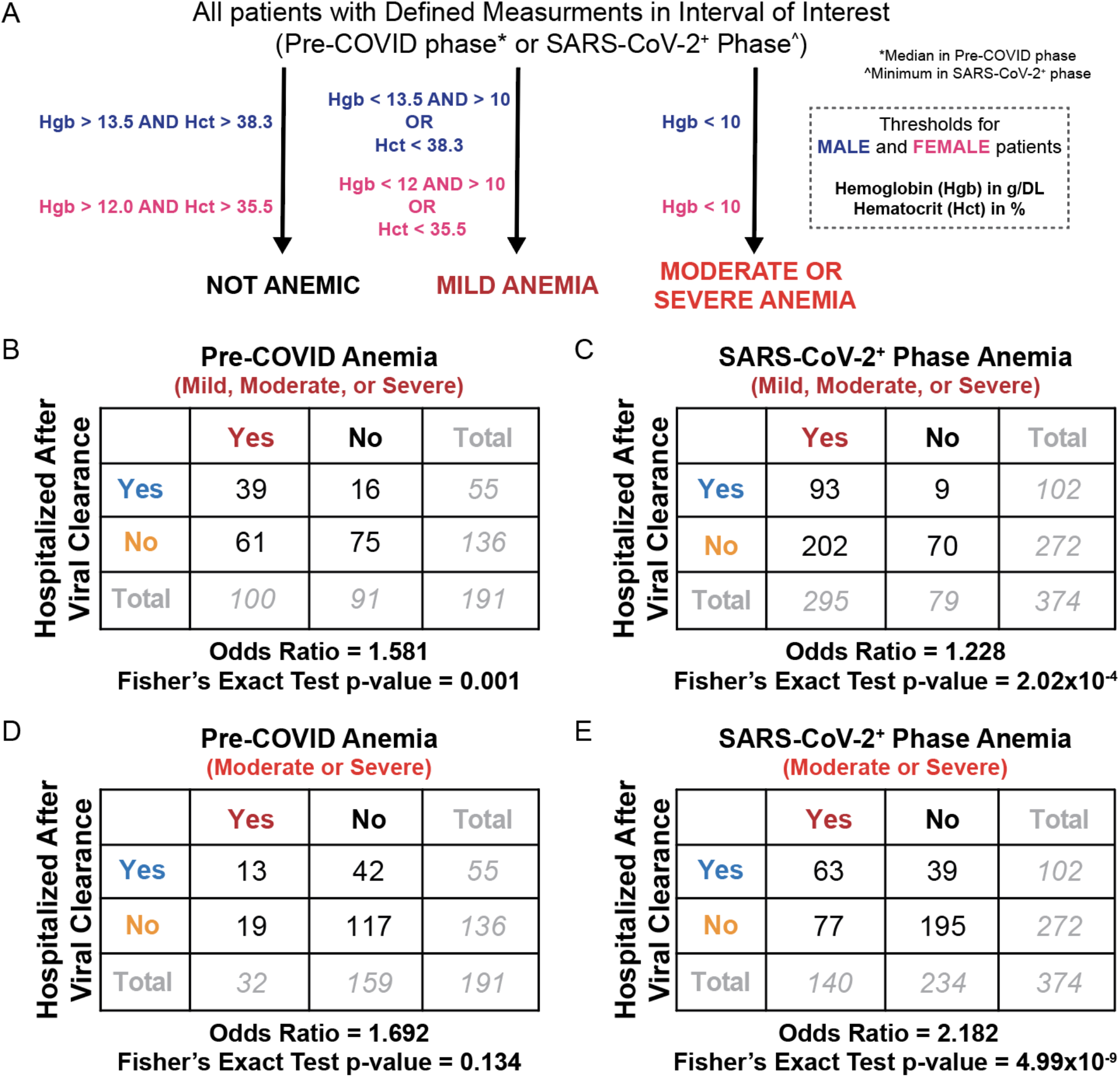
Comparison of outright anemia prevalence in the PCH and PCNH cohorts. (A) Schematic illustrating how patients are classified as having no anemia, mild anemia, or severe/moderate anemia. (B-C) Comparison of mild, moderate, or severe anemia frequency in the PCH and PCNH cohorts during the pre-COVID (n=191) and SARS-CoV-2^+^ (n=374) phases. (D-E) Comparison of moderate or severe anemia frequency in the PCH and PCNH cohorts during the pre-COVID (n=191) and SARS-CoV-2^+^ (n=374) phases. Contingency tables show the counts of patients in each intersecting category. Below each contingency table, the associated odds ratio and Fisher Exact test p-value is shown.

Similarly we assessed whether laboratory-diagnosed AKI occurs more frequently in the PCH cohort based on the creatinine-related components of the KDIGO (Kidney Disease: Improving Global Outcomes) criteria for diagnosis and staging of AKI in adults (**Figure S3A**) (Khwaja, 2012). Any stage AKI was indeed more common in the PCH cohort than the PCNH cohort in both the pre-COVID phase (30/69 [43%] vs 45/198 [23%]; OR=1.91; p=2.0×10^−3^) (**Figure S3B**) and the SARS-CoV-2^+^ phase (46/103 [45%] vs. 64/272 [24%]; OR=1.90; p=1.19×10^−4^) (**Figure S3C**). Further, stage 2+ AKI was more common in the PCH cohort during both the pre-COVID phase (21/69 [30%] vs. 21/198 [11%]; OR=2.87; p=2.24×10^−4^) and the SARS-CoV-2^+^phase (23/103 [22%] vs. 24/272 [9%]; OR=2.53; p=7.91×10^−4^) (**Figure S3D-E**). Intriguingly when split by sex, we found that this trend was driven by males, as PCH males were more likely to experience stage 2+ AKI in both the pre-COVID phase (14/37 [38%] vs. 13/109 [12%]; OR=3.17; p=0.001) and the SARS-CoV-2^+^ phase (15/53 [28%] vs. 14/158 [9%]; OR=3.19; p=9.15×10^−4^) (**Figure S3F-G**), whereas this was not true when comparing PCH and PCNH females (data not shown).

### Anemia is strongly associated with post-clearance rehospitalization independent of ICU admission status and other covariates

To test whether the previous observations were affected by potential confounding demographic or clinical covariates, we performed a series of logistic regression analyses. First we evaluated the association between post-clearance rehospitalization and the following independent variables during the pre-COVID and SARS-CoV-2^+^ phases, separately: minimum hemoglobin, maximum BUN, sex, number of blood draws, and ICU admission status (for the SARS-CoV-2^+^ phase only) (**Table 9**). The consideration of ICU admission status as a covariate was particularly important because ICU admission was more common among PCH than PCNH patients (**Table 1**). While none of these variables in the pre-COVID phase showed a significant association with rehospitalization status, minimum hemoglobin during the SARS-CoV-2^+^ phase was singularly associated with rehospitalization (β=-0.29, p=3.0×10^−5^).

**Table 9.** Logistic regression analyses to assess the association between post-viral clearance hospitalization and minimum hemoglobin, maximum BUN, or potential confounding variables during the pre-COVID and SARS-CoV-2^+^ phases. Confounding variables considered include sex, the number of blood draws in the given interval, and ICU admission status during the given interval. For each regression (row), the coefficient (β) and associated Bonferroni-adjusted p-value (p) are shown for each independent variable (column) assessed. The coefficient represents the log-odds ratio, and p-values were calculated using the log likelihood ratio test. An association between an independent variable and post clearance hospitalization is considered significant if p < 0.05 (shown in bold). The association between post-viral clearance hospitalization and ICU admission during the pre-COVID interval was not analyzed because this information was not available for our cohort prior to April 2020. Binary variables were assigned as follows: sex: 0 = female, 1 = male; ICU admission during interval: 0 = not admitted to ICU, 1 = admitted to ICU.

|  | Minimum Hemoglobin | Maximum BUN | Sex | Blood Draw Count During Interval | ICU Admission During Interval |
| --- | --- | --- | --- | --- | --- |
| **Pre-COVID Phase** | $\beta = -0.102$<br>$p = 0.547$ | $\beta = 0.012$<br>$p = 0.432$ | $\beta = 0.097$<br>$p = 0.998$ | $\beta = -0.013$<br>$p = 0.807$ | NA |
| **SARS-CoV-2<sup>+</sup> Phase** | $\beta = -0.291$<br>**$p = 3.00 \times 10^{-5}$** | $\beta = 0.006$<br>$p = 0.823$ | $\beta = -0.178$<br>$p = 0.968$ | $\beta = -0.003$<br>$p = 0.928$ | $\beta = 0.589$<br>$p = 0.161$ |

We then modified our logistic regression analysis by replacing the minimum hemoglobin and maximum BUN terms with binary labels of moderate/severe anemia and stage 2+ AKI, respectively (**Table 10**). Among the tested pre-COVID variables, only stage 2+ AKI was modestly associated with rehospitalization status (β=0.99, p=0.04). ICU admission during the SARS-CoV- 2^+^ phase was also modestly associated with post-clearance rehospitalization (β=0.70, p=0.05), suggesting that patients with more severe courses of initial COVID-19 illness are more likely to experience subsequent hospitalization. However, moderate/severe anemia during the SARS-CoV-2^+^ phase was again the most strongly associated variable with rehospitalization (β=1.16, p=6.0×10^−5^).

**Table 10.**
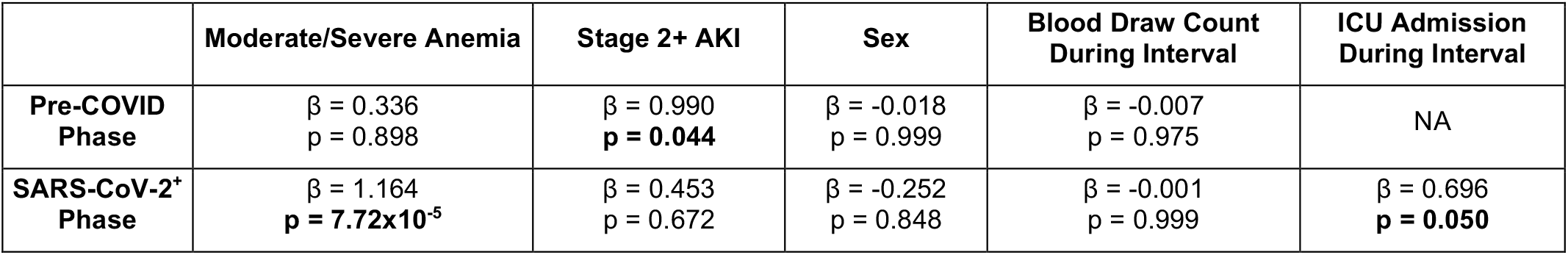
Logistic regression analyses to assess the association between post-viral clearance hospitalization and the diagnosis of moderate to severe anemia, the diagnosis of stage 2+ AKI, or potential confounding variables during the pre-COVID and SARS-CoV-2^+^ phases. Confounding variables considered include sex, the number of blood draws in the given interval, and ICU admission status during the given interval. For each regression (row), the coefficient (β) and associated Bonferroni-adjusted p-value (p) are shown for each independent variable (column) assessed. The coefficient represents the log-odds ratio, and p-values were calculated using the log likelihood ratio test. An association between an independent variable and post clearance hospitalization is considered significant if p < 0.05 (shown in bold). The association between post-viral clearance hospitalization and ICU admission during the pre-COVID interval was not analyzed because this information was not available for our cohort prior to April 2020. Binary variables were assigned as follows: moderate/severe anemia: 0 = no anemia, 1 = anemia; sex: 0 = female, 1 = male; ICU admission during interval: 0 = not admitted to ICU, 1 = admitted to ICU.

Finally we performed a split cohort subanalysis to specifically test whether the robust association between anemia and rehospitalization status applied to both patients who were and were not admitted to the ICU during their index infection. Indeed, the rate of moderate/severe anemia was significantly higher in PCH patients than PCNH patients when considering only those were were admitted to the ICU (59% [38/64] vs. 19% [10/53], OR=2.10, p=1.09×10^−5^) or only those who were not admitted to the ICU (33% [25/76] vs. 16% [29/181], OR=1.84, p=4.0×10^−3^) (**Figures S4A-B**).

## Discussion

Approximately one year after the first confirmed case, the COVID-19 pandemic continues to ravage communities across the globe. While efforts early in the pandemic rightly focused on the acute lung inflammation caused by SARS-CoV-2, the subsequent realization that COVID-19 may have more lasting effects has mandated a better understanding of factors that predispose patients to experience long-term COVID-19 related complications. We have previously sought to address this knowledge gap using state-of-the-art NLP models deployed on a complete EHR system (Pawlowski *et al*., 2020), and here we have expanded this effort to include the longitudinal analysis of laboratory measurements both prior to COVID-19 diagnosis and during active SARS-CoV-2 infection.

This lab test analysis shows that anemia and renal function in the pre-COVID and SARS-CoV-2^+^ phases are associated with the risk of post viral clearance hospitalization. Our logistic regression analyses suggest that AKI and static renal lab measurements are not independently associated with rehospitalization, with ICU admission during COVID-19 infection representing a likely confounding factor contributing to the observed trends. Indeed, AKI has previously been reported as a common morbidity in ICU patients (Mohsenin, 2017) and was observed frequently among ICU admitted COVID-19 patients in our cohort, with 78% (94/121) and 39% (47/121) of ICU admitted patients experiencing stage 1+ and stage 2+ AKI, respectively. On the other hand, hemoglobin levels and the outright diagnosis of moderate or severe anemia are robustly associated with post-clearance rehospitalization independent of sex, number of blood draws, and ICU admission status.

While the pathophysiologic foundations for these associations are not yet clear, the findings do merit consideration in the context of COVID-19 clinical care. Indeed, pre-existing conditions are already integrated in the clinical decision-making algorithms around COVID-19 as the Center for Diseases Control (CDC) has designated various chronic conditions as risk factors for severe COVID-19 infection (e.g. cancer, chronic kidney disease, chronic obstructive pulmonary disease, and cardiovascular diseases such as heart disease, obesity, and diabetes) (CDC, 2020). However, there is much less known regarding factors or conditions that place people at risk for subsequent complications such as rehospitalization after viral clearance. Once identified, such factors and conditions should similarly be incorporated into the clinical decision-making process when treating COVID-19 patients.

Our finding that lower hemoglobin and hematocrit levels, and the outright diagnosis of moderate or severe anemia, prior to or during active SARS-CoV-2 infection is associated with post viral clearance hospitalization has not been previously reported. And while sickle cell disease is considered a risk factor for severe COVID-19, anemia itself is not considered to be such a risk factor. While a causal role for anemia in post clearance hospitalization is certainly not established by this analysis, the robust association warrants further studies to determine the whether anemia mitigating therapies (e.g., vitamin or mineral supplementation, erythropoietin administration, or blood transfusion) engender long-term benefits in select COVID-19 patients. Further, even if mitigation of anemia does not impact subsequent hospitalization, these links between anemia and COVID-19 are interesting in light of several previous lines of research.

First, several groups have reported an association between blood groups and susceptibility to or severity of COVID-19 infection (Latz *et al*., 2020, Ellinghaus *et al*., 2020). Specifically, it has been suggested that individuals with type O blood may be at lower risk for contracting COVID-19 or experiencing respiratory failure in the context of COVID-19. Whether this association reflects a direct or indirect interaction between SARS-CoV-2 and erythrocytes is not known, but it could certainly be relevant to pursue whether blood type is also associated with the occurrence or severity of anemia in the setting of COVID-19.

Second, fatigue has been commonly reported as both an acute symptom and a lasting effect of COVID-19 (Wagner *et al*., 2020, Pascarella *et al*., 2020, Townsend *et al*., 2020), but the mechanisms underlying this phenotype have not been established. It is worth noting that 295 of the 374 (79%) hospitalized COVID-19 patients in this study had at least mild anemia during their SARS-CoV-2^+^ phase, and 140 of the 374 (37%) patients had moderate or severe anemia (defined as hemoglobin < 10 g/dL) during this interval. It would be worthwhile to perform a longitudinal follow-up on these patients to determine whether they continue to experience anemia in the months following SARS-CoV-2 clearance, and whether the presence of such a post-COVID anemia is associated with reports of fatigue.

Along with our previous analysis (Pawlowski *et al*., 2020), this study illustrates the value of deploying sophisticated platforms across EHR systems that enable the integrated analysis of diverse data types including sentiment-laden text and laboratory test measurements. Taken together, these studies provide the first example of leveraging augmented curation methods to first identify phenotypes that distinguish defined clinical cohorts and to then cross check these phenotypic associations through a hypothesis-driven analysis of the most relevant lab tests. This framework can be effectively scaled for other clinical research efforts not only in COVID-19 but also in any other disease areas of interest.

### Limitations of the Study

This study has several important limitations to consider. First, this analysis considers only patients within one EHR system; while this system does contain patient data from multiple sites of clinical care in distinct geographic locations (Minnesota, Arizona, Florida), there are still likely underlying biases in important factors such as patient demographics and tendencies around the ordering of laboratory tests by clinicians. Such biases would prevent the studied cohort and their associated data points from serving as true representative samples of all COVID-19 patients. Second, the analyzed cohort was relatively small (n = 382) as most patients diagnosed with COVID-19 do not subsequently receive two confirmatory negative PCR tests, and only a subset of these 382 patients possessed data for each lab test of interest. Finally, the definition of the SARS-CoV-2^+^ window is imperfect as the true date of viral clearance for a given patient would likely precede their first negative PCR test by an unknown amount of time.

### Resource Availability

After publication, the data will be made available to others upon reasonable requests to the corresponding author. A proposal with a detailed description of study objectives and the statistical analysis plan will be needed for evaluation of the reasonability of requests. Deidentified data will be provided after approval from the corresponding author and Mayo Clinic.

## Methods

### Study design

This was a case-control study. The primary outcome was rehospitalization status within 30 days of PCR-confirmed SARS-CoV-2 clearance. The exposure variables were anemia and kidney dysfunction as assessed through selected lab measurements detailed below.

### Selection of study participants

Cases and controls were selected from a cohort of 66,689 patients who presented to the Mayo Clinic Health System (including tertiary medical centers in Minnesota, Arizona, and Florida) and received at least one positive SARS-CoV-2 PCR test between the start of the COVID-19 pandemic and December 12, 2020 (see **Figure 1B**). Post clearance hospitalized (“PCH”) cases (n=104) were defined as patients who were hospitalized for COVID-19, had two documented negative SARS-CoV-2 PCR tests following their last positive test result, and were subsequently admitted to the hospital within 30 days of clearance. Post clearance non-hospitalized (“PCNH”) controls (n=278) were defined as those who were hospitalized for COVID-19, had two documented negative SARS-CoV-2 PCR tests following their last positive test result, and were not hospitalized within 30 days of clearance. Demographic and clinical features of the PCH and PCNH cohorts are summarized in **Table 1**.

### Definition of the considered time intervals: pre-COVID phase and SARS-CoV-2^+^ phase

Laboratory results were assessed (1) during the year prior to COVID-19 diagnosis, referred to throughout this manuscript as the “pre-COVID phase” and (2) during the period in which a patient was positive for SARS-CoV-2 by PCR, referred to throughout this manuscript as the “SARS-CoV-2^+^ phase.” COVID-19 diagnosis was conferred by a positive SARS-CoV-2 PCR test, and clearance was defined as two consecutive negative SARS-CoV-2 PCR tests occurring after a positive test. The estimated viral clearance date was taken as the date of the first negative PCR test in this sequence of two consecutive negative tests.

### Selection and summarization of laboratory measurements

The primary exposure variables were anemia and AKI. The selected laboratory measurements related to anemia included hemoglobin and hematocrit, and laboratory measurements related to AKI included serum creatinine, serum blood urea nitrogen (BUN), and estimated glomerular filtration rate (eGFR). The majority of eGFR measurements (∼96%) were estimated by creatinine; these tests had a maximum recorded value of 90 mL/min/BSA, which corresponds to the lower limit of normal. The remaining 4% of eGFR measurements were estimated by cystatin C levels; for these tests, a value above 90 mL/min/BSA was possible and was indeed recorded in 5 of 31 cases.

For a given lab test, we considered the median, maximum, and minimum measurements for each patient during the specified time windows (i.e. the pre-COVID and SARS-CoV-2^+^ phases). Histograms showing the number of measurements per patient in each time period for the selected tests are shown in **Figures S5-S6**. Given the directionality of these tests (i.e. anemia is defined by low hemoglobin and hematocrit, while kidney dysfunction is characterized by increases in serum creatinine and BUN but a decrease in eGFR), we were primarily interested in comparing the patient-level minimum values of hemoglobin, hematocrit, and eGFR, and maximum values of serum creatinine and BUN in each time period.

### Consideration of potential confounding variables

As shown in **Table 1**, there were no statistically significant differences between these groups in age, relative cleared date (defined as the time to the first negative SARS-CoV-2 PCR test in a series of two consecutive negative tests after the last positive test), race, ethnicity, or sex. However, we did note that a higher fraction of PCNH cases were male as compared to PCH counterparts (58% vs. 52%). This potential confounding factor was addressed by performing (1) sex-split subgroup analyses (**Tables 5-8**) and (2) multivariate logistic regression (**Tables 9-10**; see *Statistics* below).

Although hospitalization during index infection was required for inclusion in both the PCH and PCNH cohorts, this criterion does not necessarily ensure comparable severities of index infection. To better assess potential differences in index infection severity, we compared the rates of ICU admission and found this to be significantly higher in the PCH cohort compared to the PCNH cohort (48/104 [46%] vs. 73/278 [26%], p=4.0×10^−3^; **Table 1**). Further, patients admitted to the ICU during index infection had slightly lower hemoglobin measurements during the SARS-CoV-2^+^ phase than patients not admitted to the ICU (cohen’s D = -0.27, p=0.01; **Figure S7**). Thus, we considered ICU admission as a potential confounding factor in our analyses. We addressed this by performing (1) subgroup analyses to determine whether differences between the PCH and PCNH cohorts were observed both in patients who were and were not admitted to the ICU (**Figure S4**) and (2) multivariate logistic regression (**Tables 9-10**; see *Statistics* below).

We observed that patients in the PCH cohort were more likely to experience anemia in both the pre-COVID and SARS-CoV-2^+^ phases than patients in the PCNH cohort. Because hospitalized patients can experience anemia due to repeated blood draws for laboratory testing, we also considered the number of blood draws per patient as a potential confounding variable. To address this, we performed multivariate logistic regression (**Tables 9-10**; see *Statistics* below).

### Classification of patients using clinical diagnostic criteria for anemia and AKI

We classified patients in a binary fashion for each time window based on whether their lab tests were consistent with the clinical diagnosis of anemia or acute kidney injury. Classifications were defined according to the Mayo Clinic reference ranges for anemia and the KDIGO (Kidney Disease: Improving Global Outcomes) criteria for AKI (Khwaja, 2012) as follows (see also **Figure 5A** and **Figure S3A**):

- *Anemia (mild, moderate, or severe)*: for males, median hemoglobin < 13.5 g/dL or median hematocrit < 38.3%. For females, median hemoglobin < 12.0 g/dL or median hematocrit < 35.5%.
- *Anemia (moderate or severe)*: for both males and females, median hemoglobin < 10.0 g/dL.
- *Acute kidney injury (stage 1, 2, or 3)*: increase in serum creatinine by ≥0.3 mg/dL within 48 hours or an increase in serum creatinine to ≥1.5x the baseline value which is known or assumed to have occurred in the prior 7 days. The baseline was defined as the minimum value among all serum creatinine tests for the given patient in the prior 7 days.
- *Acute kidney injury (stage 2 or 3)*: increase in serum creatinine to ≥2x the baseline value which is known or assumed to have occurred in the prior 7 days, or a serum creatinine value of ≥4 mg/dL. The baseline was defined as the minimum value among all serum creatinine tests for the given patient in the prior 7 days.

### Quantification of number of blood draws per patient

To test whether trends in anemia-related measurements could be explained by differences in the number of blood draws received in the pre-COVID phase or SARS-CoV-2^+^ phase, we counted the number of blood draws in these time intervals for each patient. All tests with a documented source of “Blood”, “Plasma”, or “Serum” were first collected for each patient. For a given patient on a given day, we then took the count of the most frequently obtained test as the number of blood draws for that patient on that day. For example, if the record for Patient *P* on Day *D* contained 5 serum sodium measurements, 3 hemoglobin measurements, and 1 plasma IL-6 measurement, then we inferred that Patient *P* received 5 blood draws on Day *D*.

### Statistics

Laboratory values were assessed within each time interval as patient-wise medians, minima, or maxima. To perform statistical comparisons between the PCH and PCNH cohorts, one-sided Mann-Whitney U-tests and Cohen’s D were applied to continuous outcome measures, generating a *p*-value and an effect size measurement. The distribution of patient-wise median, minimum, and maximum values obtained for each laboratory measurement among this cohort were assessed with a Kolmogorov-Smirnov (KS) Test of Normality (**Figures S8-S10**). As these measurements did not follow a normal distribution (KS Test p-value < 0.05), the non-parametric Mann-Whitney U test was chosen for statistical comparisons. A one-sided test was used because these comparisons were performed as follow-up to our previous EHR-based analysis which found a higher prevalence of anemia and kidney injury in the PCH cohort (Pawlowski *et al*., 2020), providing a pre-supposed direction of change for each tested laboratory measurement. For each set of comparisons performed, p-values were corrected using a Benjamini-Hochberg (BH) correction for multiple hypothesis testing. Differences were considered statistically significant and biologically relevant if the BH-corrected p-value was ≤ 0.05 and the cohen’s D magnitude was ≥ 0.4. Fisher exact tests were applied to categorical outcome measures, generating a *p*-value and an Odds Ratio. All of the tests described above were applied using the SciPy package (Virtanen *et al*., 2020) in Python (version 3.5).

To address potential confounding variables that may be related to the observed trends in laboratory measurements, we performed multivariate logistic regressions for both the pre-COVID and SARS-CoV-2^+^ phases (**Tables 9-10**). For each regression, the binary dependent variable was defined as post viral clearance hospitalization status (i.e. assignment to the PCH cohort vs. PCNH cohort), and the independent variables included one anemia metric and one renal function metric along with sex (binary), number of blood draws (continuous) in the given time interval, and ICU admission status during that time interval (binary). Stated explicitly, the logistic regression equation was as follows: *log(P*_*PCH*_*/(1 - P*_*PCH*_*)) = β*_*0*_ *+ β*_*1*_*\*(Anemia Metric) + β*_*2*_*\*(Renal Function Metric) + β*_*3*_*\*(Sex) + β*_*4*_*\*(Blood Draw Count) + β*_*5*_*\*(ICU Admission Status)*. In one set of logistic regressions, the anemia and renal function metrics employed were minimum hemoglobin (continuous) and maximum BUN (continuous), respectively (**Table 9**). In another set, the anemia and renal function metrics employed were diagnosis status for moderate or severe anemia (binary) and diagnosis status for stage 2+ AKI (binary), respectively (**Table 10**). Binary variables were assigned as follows: anemia: 0 = no anemia or mild anemia, 1 = moderate or severe anemia; AKI: 0 = no AKI or stage 1 AKI, 1 = stage 2+ AKI; sex: 0 = female, 1 = male; ICU admission during interval: 0 = not admitted to ICU, 1 = admitted to ICU. Of note, data regarding ICU admission status was not available prior to April 2020, so this feature was omitted from the pre-COVID regression analyses.

For each time interval, two regressions were performed: one using the continuous metrics for anemia and AKI, and one using the binarized metrics for anemia and AKI. We performed separate regressions for the continuous and binarized anemia and AKI terms rather than including all terms in one model because these metrics are strongly correlated with each other, and multicollinearity of independent variables can negatively impact the estimation of logistic regression coefficients. Each regression yielded a coefficient (log odds ratios) and p-value (calculated using the log likelihood ratio test) for each independent variable. P-values were adjusted within the output of each model using the Bonferroni correction: p_Bonf_ = 1-(1-p)^*n*^. Here, *n* corresponds to the number of independent variables tested (4 for pre-COVID models, 5 for SARS-CoV-2^+^ models). All regressions were performed using the Statsmodels package in python (Seabold and Perktold, 2010).

### Study Approval

This retrospective study was reviewed and approved by the Mayo Clinic Institutional Review Board (IRB 20-003278) as a minimal risk study. Subjects were excluded if they did not have a research authorization on file.

## Acknowledgements

We thank Murali Aravamudan for his thoughtful review and feedback on this manuscript. We also thank Andrew Danielsen, Jason Ross, Jeff Anderson, and Sankar Ardhanari for their support that enabled the rapid completion of this study. The authors acknowledge funding from nference for this study.

## Author Contributions

PL, ER, AV, RM, ADB, and VS contributed to the study design and methodology. ER, GB, and JCO were responsible for data curation. ER performed the formal statistical analyses, which were reviewed by PL, AV, RM, and VS. PL drafted the manuscript with inputs from ER. AV, RM, JCO, ADB, WH, JH, and VS provided critical revisions, which were incorporated into the final manuscript by PL. All authors approved the submitted version of the manuscript.

## Supplemental Information

**Figure S1.**
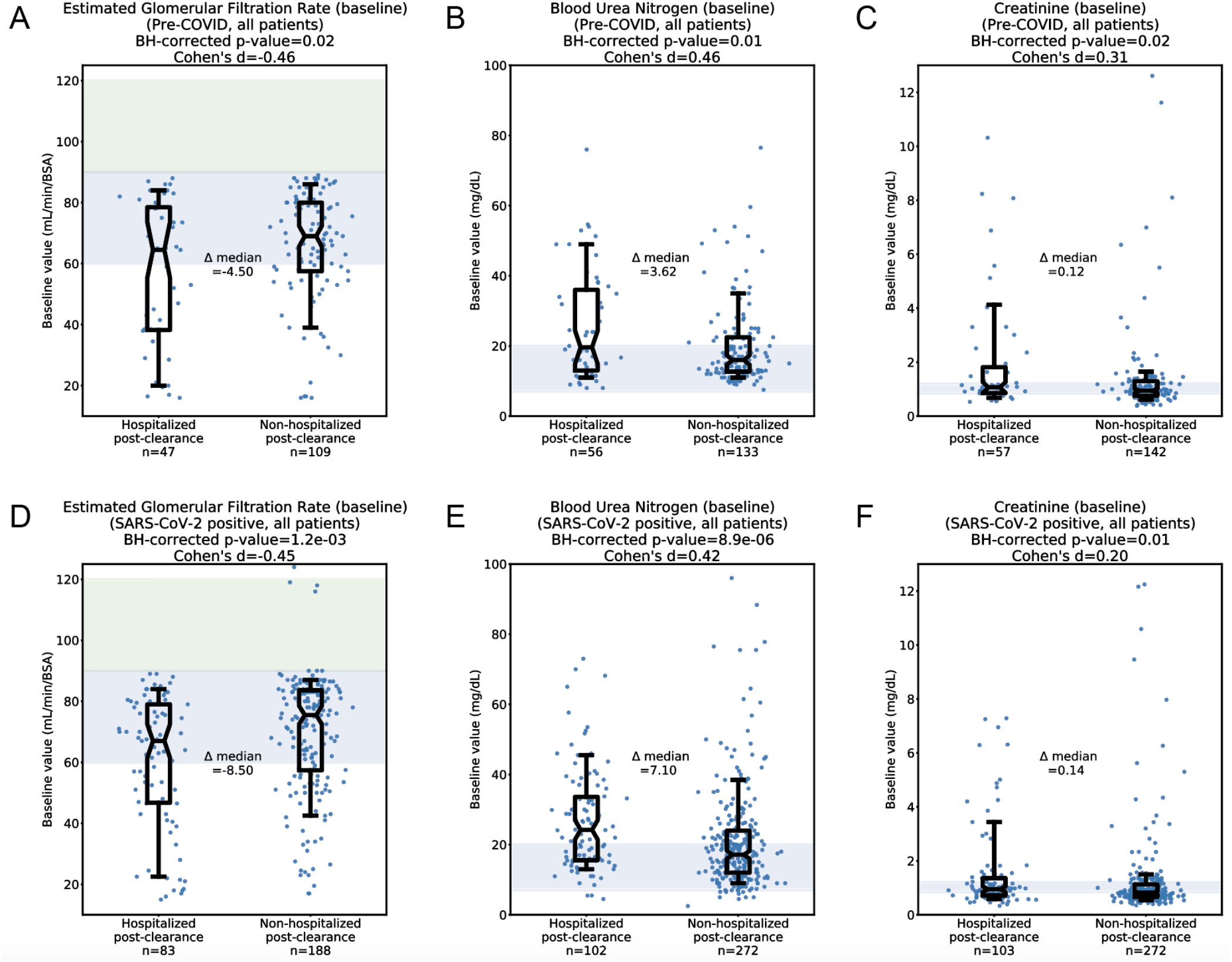
Comparison of median values for renal function tests during the pre-COVID and SARS-CoV-2^+^ phases. (A) Pre-COVID median eGFR in the PCH (n=47) and PCNH (n=109) cohorts. (B) Pre-COVID median BUN in the PCH (n=56) and PCNH (n=133) cohorts. (C) Pre- COVID median serum creatinine in the PCH (n=57) and PCNH (n=142) cohorts. (D) SARS-CoV- 2^+^ median eGFR in the PCH (n=83) and PCNH (n=188) cohorts. (E) SARS-CoV-2^+^ median BUN in the PCH (n=102) and PCNH (n=272) cohorts. (F) SARS-CoV-2^+^ median serum creatinine in the PCH (n=103) and PCNH (n=272) cohorts. Shaded regions correspond to normal ranges for each test. For eGFR, the blue shading (60-90 mL/min/BSA) indicates moderately reduced levels which can be considered normal in older patients, while the green shading (>90 mL/min/BSA) indicates the normal range for younger patients. Normal ranges shown for BUN and serum creatinine are 7-20 mg/dL and 0.84-1.21 mg/dL, respectively. For each comparison, statistics shown include the number of patients analyzed, Cohen’s D, BH-corrected Mann Whitney U test p-value, and the difference of medians between the two cohorts. Box and whisker plots depict median and IQR along with the 10th and 90th percentiles.

**Figure S2.**
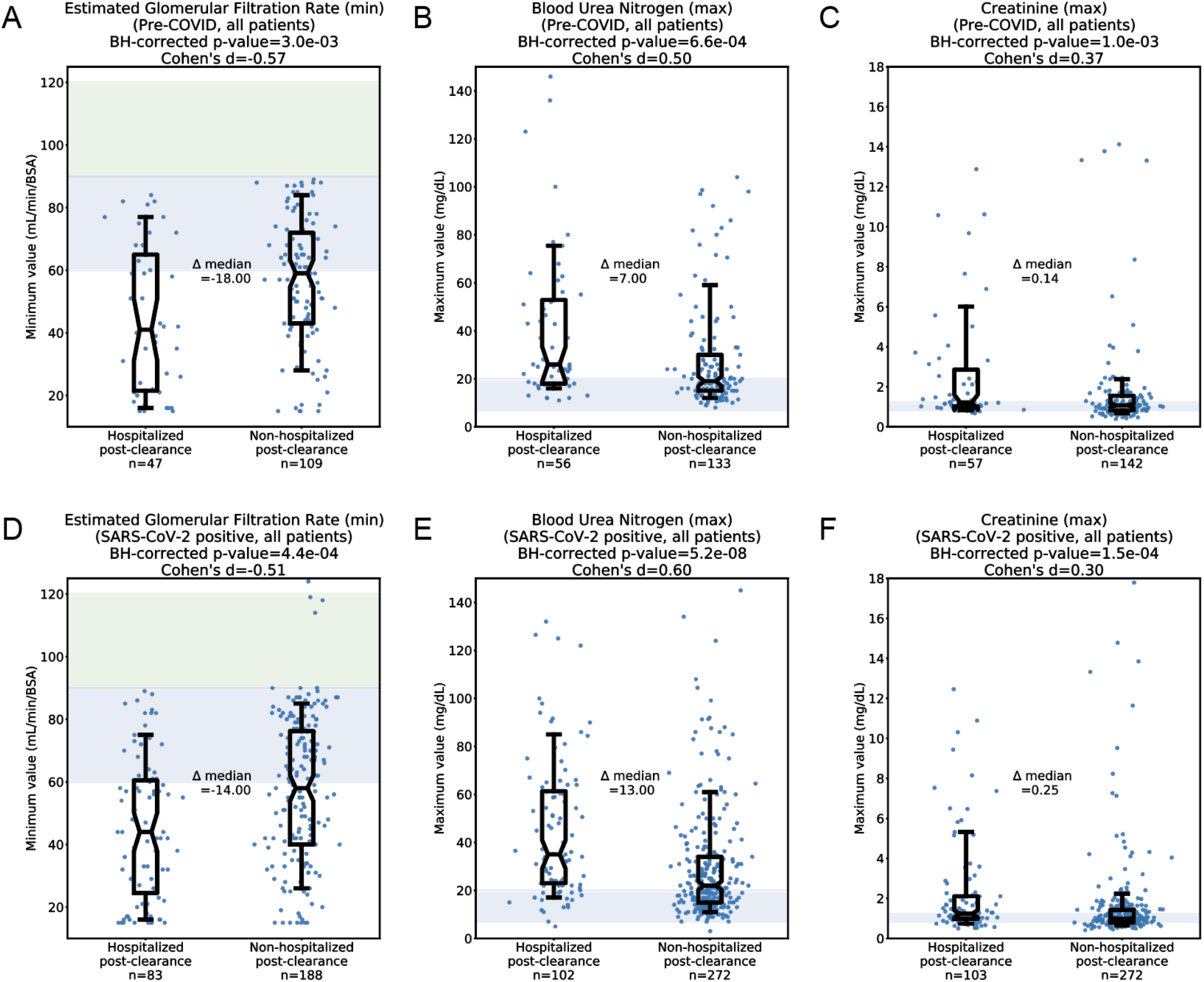
Comparison of “extreme” (minimum or maximum) values for renal function tests during the pre-COVID and SARS-CoV-2^+^ phases. (A) Pre-COVID minimum eGFR in the PCH (n=47) and PCNH (n=109) cohorts. (B) Pre-COVID maximum BUN in the PCH (n=56) and PCNH (n=133) cohorts. (C) Pre-COVID maximum serum creatinine in the PCH (n=57) and PCNH (n=142) cohorts. (D) SARS-CoV-2^+^ minimum eGFR in the PCH (n=83) and PCNH (n=188) cohorts. (E) SARS-CoV-2^+^ maximum BUN in the PCH (n=102) and PCNH (n=272) cohorts. (F) SARS-CoV- 2^+^ maximum serum creatinine in the PCH (n=103) and PCNH (n=272) cohorts. Shaded regions correspond to normal ranges for each test. For eGFR, the blue shading (60-90 mL/min/BSA) indicates moderately reduced levels which can be considered normal in older patients, while the green shading (>90 mL/min/BSA) indicates the normal range for younger patients. Normal ranges shown for BUN and serum creatinine are 7-20 mg/dL and 0.84-1.21 mg/dL, respectively. For each comparison, statistics shown include the number of patients analyzed, Cohen’s D, BH-corrected Mann Whitney U test p-value, and the difference of medians between the two cohorts. Box and whisker plots depict median and IQR along with the 10th and 90th percentiles.

**Figure S3.**
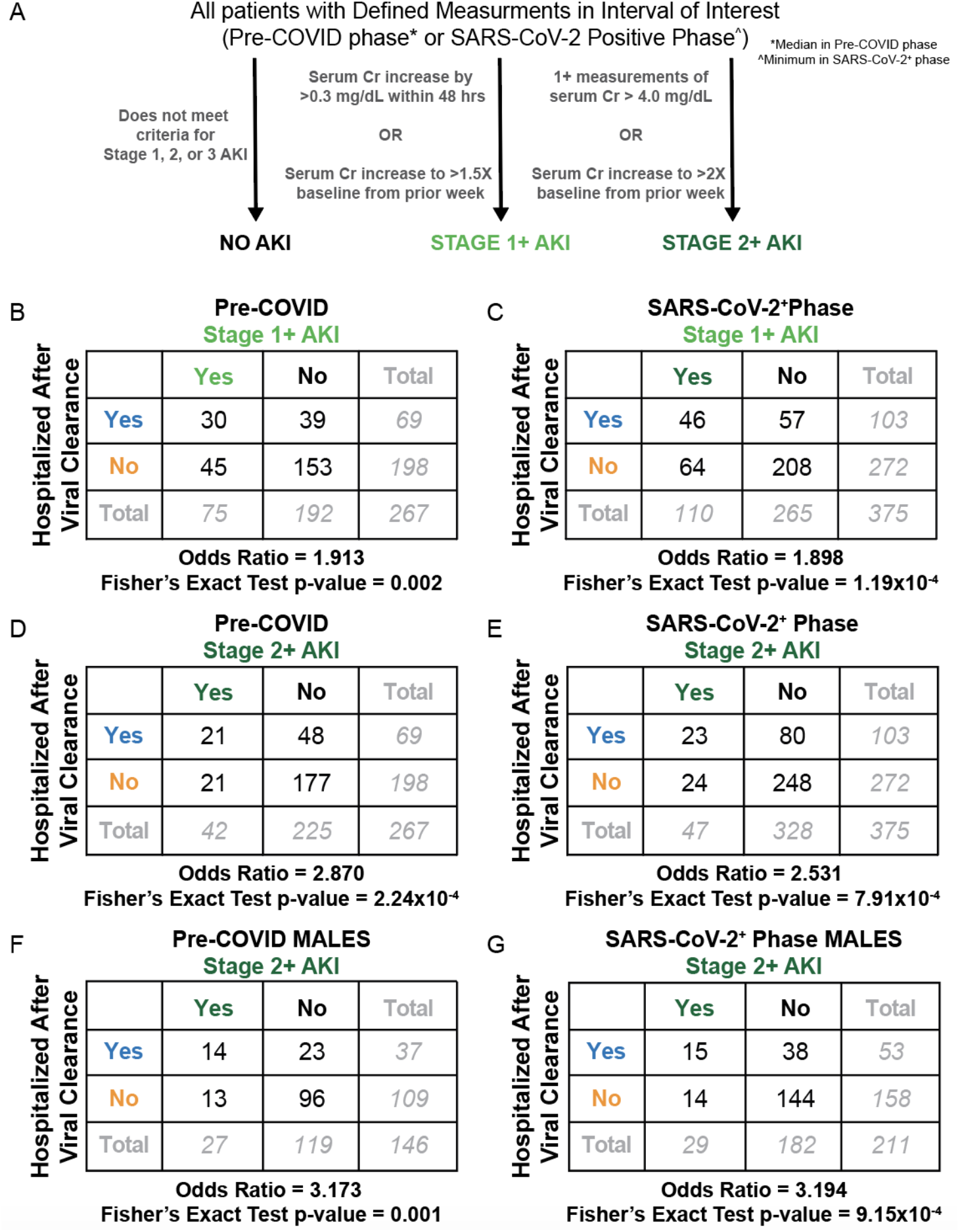
Comparison of AKI frequency in the PCH and PCNH cohorts during the pre-COVID and SARS-CoV-2^+^ phases. (A) Schematic illustrating how patients are classified as having no AKI, stage 1 AKI, or stage 2/3 AKI, based on the KDIGO criteria. (B) Association between pre- COVID AKI (any stage) and post viral clearance hospitalization status in all patients (n=267). (C) Association between SARS-CoV-2^+^ phase AKI and post viral clearance hospitalization status in all patients (n=375). (D) Association between pre-COVID Stage 2 or 3 AKI and post viral clearance hospitalization status in all patients (n=267). (E) Association between SARS-CoV-2^+^ phase Stage 2 or 3 AKI and post viral clearance hospitalization status in all patients (n=375). (F) Association between pre-COVID Stage 2 or 3 AKI and post viral clearance hospitalization status in only male patients (n=146). (F) Association between SARS-CoV-2^+^ phase Stage 2 or 3 AKI and post viral clearance hospitalization status in only male patients (n=211). Contingency tables show the counts of patients in each intersecting category. Below each contingency table, the associated odds ratio and Fisher Exact test p-value is shown.

**Figure S4.**
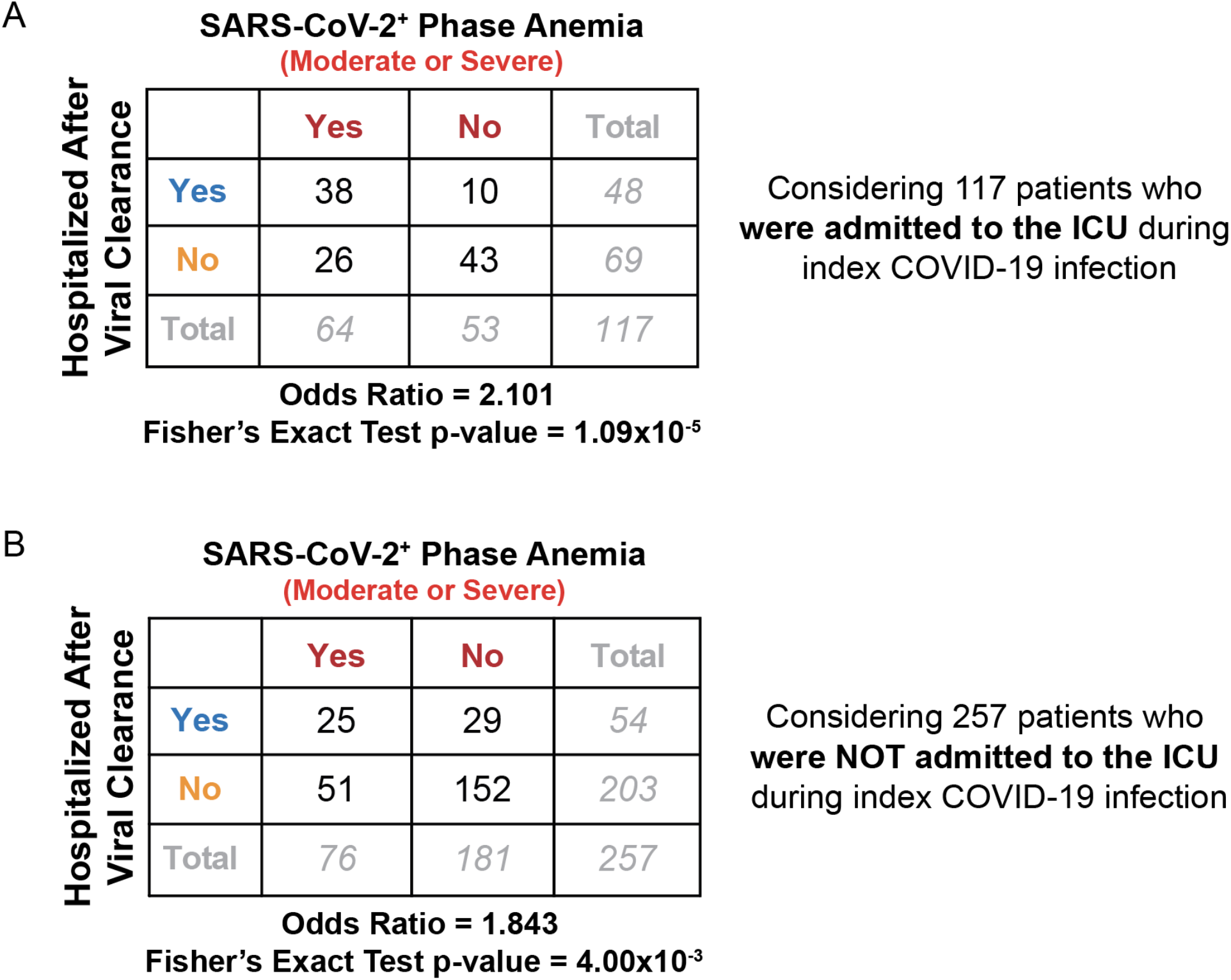
Comparison of rates of moderate and severe anemia in the PCH versus PCNH cohorts split by index infection ICU admission status. (A) Rates of moderate or severe anemia in patients who were admitted to the ICU during their index COVID-19 infection (n=117). (B) Rates of moderate or severe anemia in patients who were not admitted to the ICU during their index COVID-19 infection (n=257). Contingency tables show the counts of patients in each intersecting category. Below each contingency table, the associated odds ratio and Fisher Exact test p-value is shown. Regardless of ICU admission status, patients who experienced moderate or severe anemia during the SARS-CoV-2^+^ phase were more likely to be hospitalized after viral clearance.

**Figure S5.**
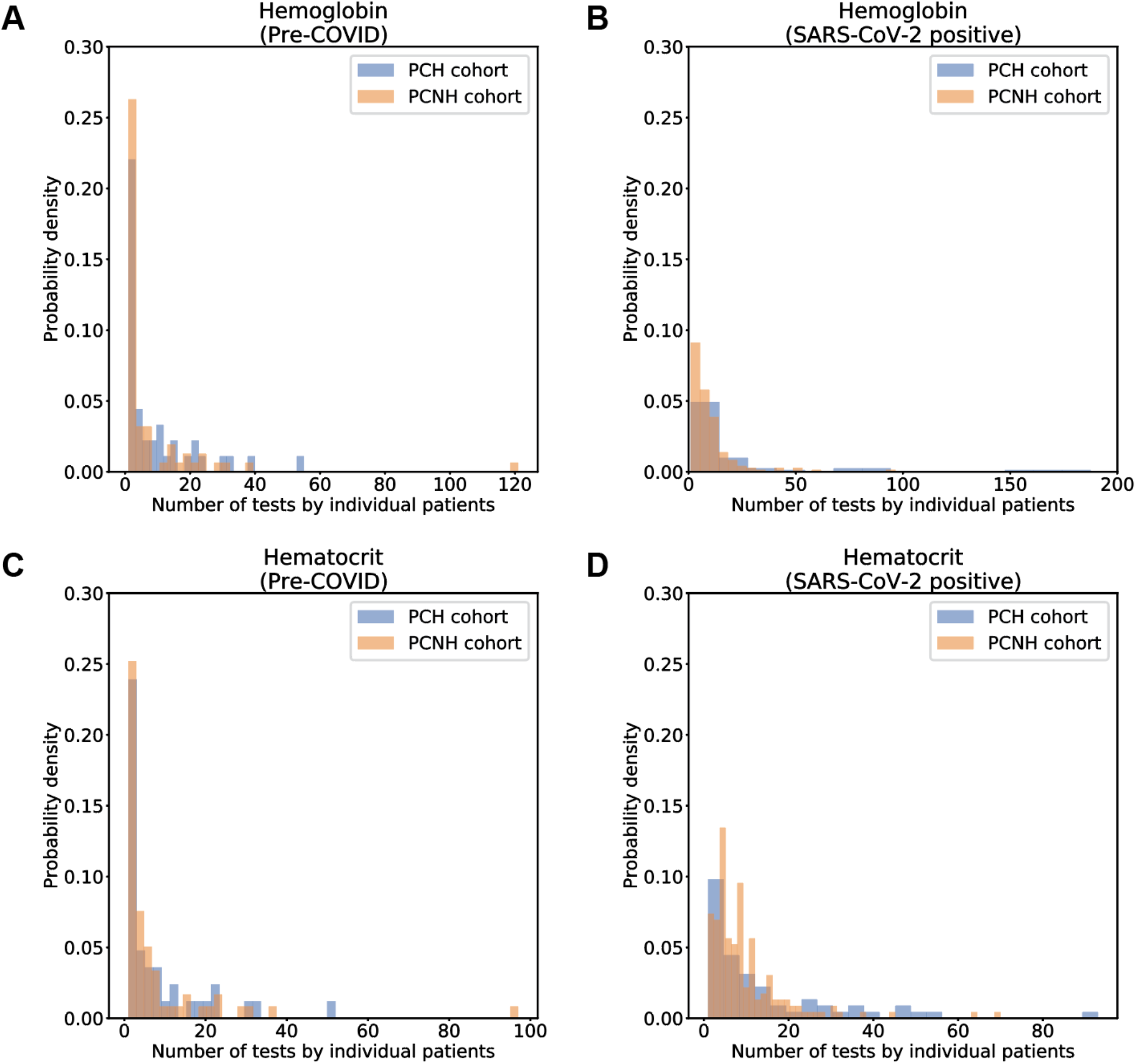
Histograms depicting the number of measurements per patient for the selected set of anemia-related lab tests in the pre-COVID and SARS-CoV-2^+^ intervals. (A) Number of hemoglobin tests per patient in the pre-COVID phase. (B) Number of hemoglobin tests per patient in the SARS-CoV-2^+^ phase. (C) Number of hematocrit tests per patient in the pre-COVID phase. (D) Number of hematocrit tests per patient in the SARS-CoV-2^+^ phase. Counts are shown separately for the PCH (blue) and PCNH (orange) cohorts.

**Figure S6.**
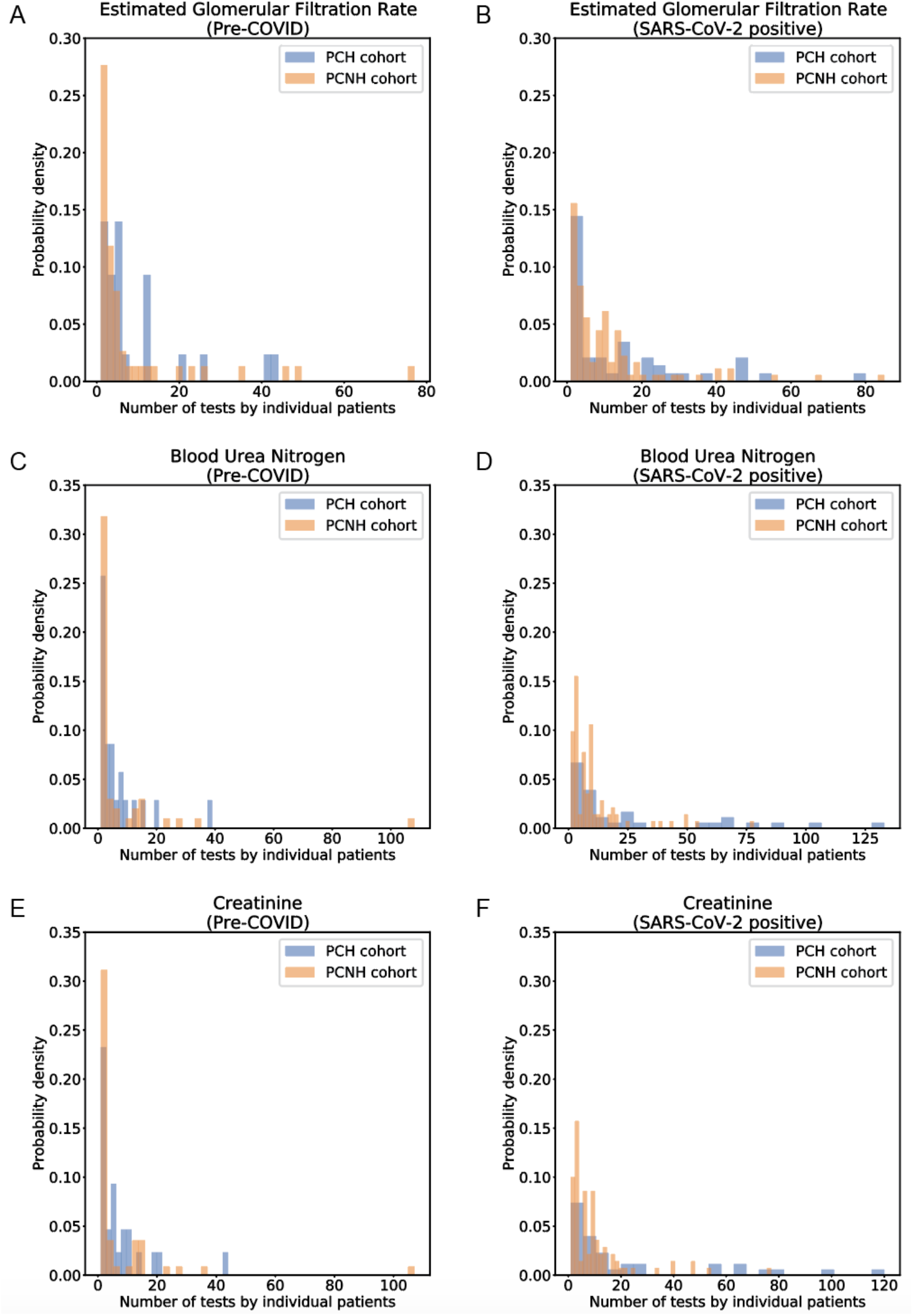
Histograms depicting the number of measurements per patient for the selected set of renal function tests in the pre-COVID and SARS-CoV-2^+^ intervals. (A) Number of eGFR tests per patient in the pre-COVID phase. (B) Number of eGFR tests per patient in the SARS-CoV-2^+^ phase. (C) Number of BUN tests per patient in the pre-COVID phase. (D) Number of BUN tests per patient in the SARS-CoV-2^+^ phase. (C) Number of serum creatinine tests per patient in the pre-COVID phase. (D) Number of serum creatinine tests per patient in the SARS-CoV-2^+^ phase. Counts are shown separately for the PCH (blue) and PCNH (orange) cohorts.

**Figure S7.**
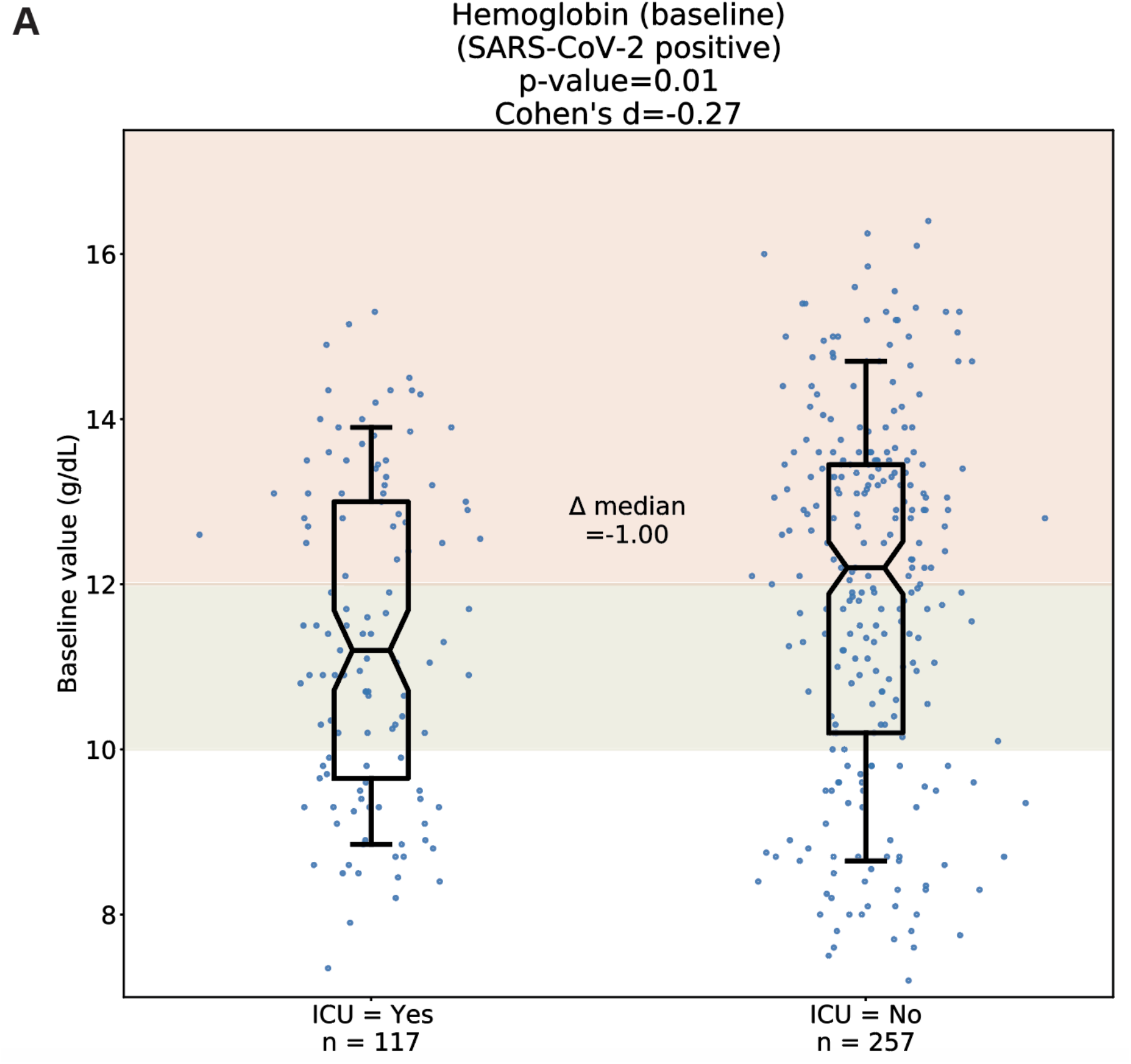
Comparison of hemoglobin levels in patients based on ICU admission status during COVID-19 index infection. Each dot corresponds to the median (“baseline”) hemoglobin level of an individual patient during the SARS-CoV-2^+^ phase. The 374 patients with hemoglobin measurements in the SARS-CoV-2^+^ phase (out of 382 total patients) were divided based on whether they were admitted to the ICU during their index infection. Red shading indicates normal ranges for hemoglobin and hematocrit; as these ranges are lower for females than males, the shaded range here spans from the lower limit of normal for females (12 g/dL hemoglobin, 35.5% hematocrit) to the upper limit of normal for males (17.5 g/dL hemoglobin, 48.6% hematocrit). Green shading indicates mild anemia, defined as a hemoglobin level greater than 10 g/dL and less than the sex-dependent lower limit of normal. Statistics shown include the number of patients per group, Cohen’s D, Mann Whitney U test p-value, and the difference of medians between the two cohorts. Box and whisker plots depict median and IQR along with the 10th and 90th percentiles.

**Figure S8.**
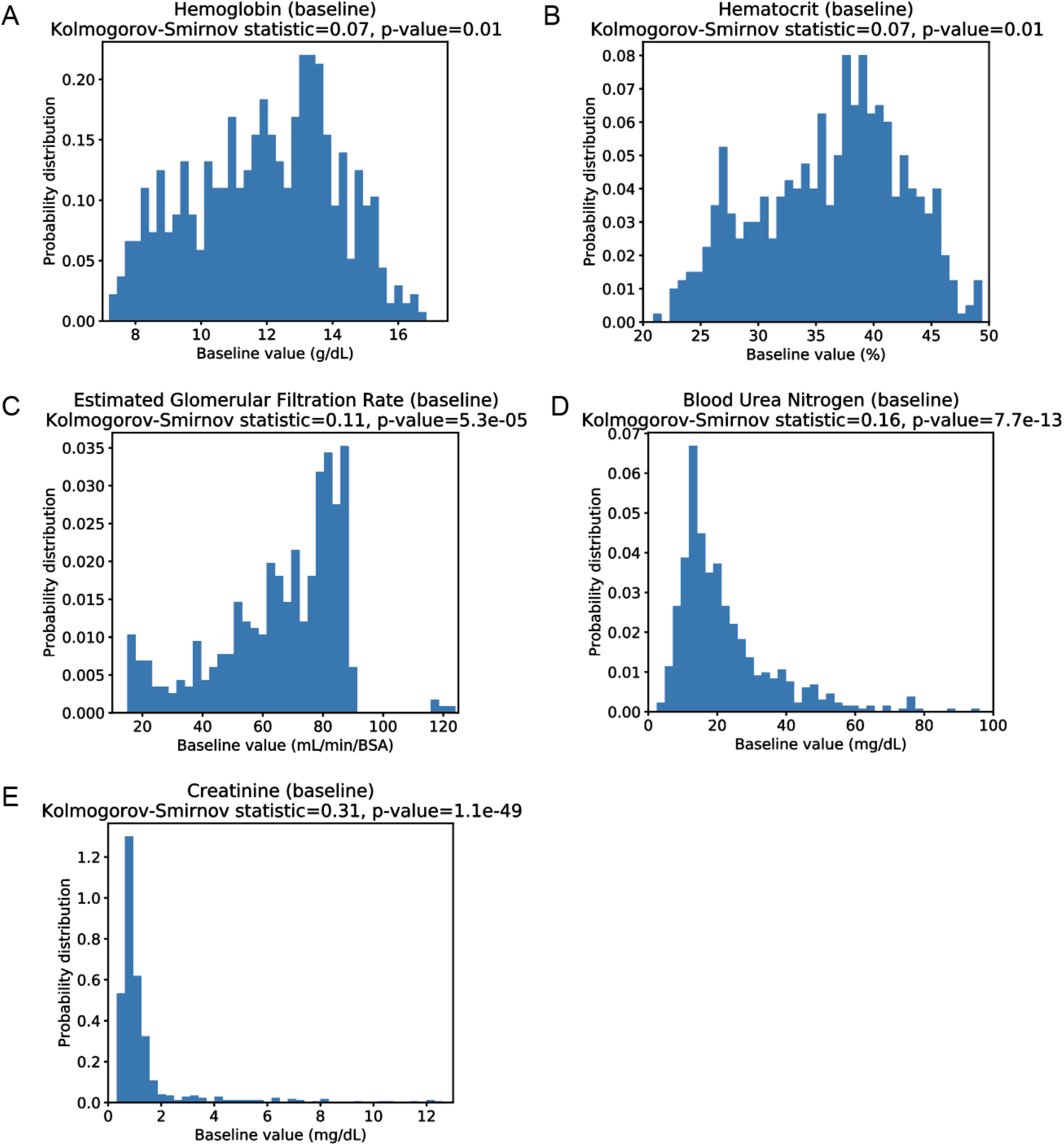
Distributions and normality tests of patient-wise median values for all considered lab tests. Lab tests evaluated include (A) hemoglobin, (B) hematocrit, (C) eGFR, (D) BUN, and (E) serum creatinine. Above each plot, the Kolmogorov-Smirnov statistic and associated p-value are shown; a p-value below 0.05 indicates that the data do not follow a normal distribution.

**Figure S9.**
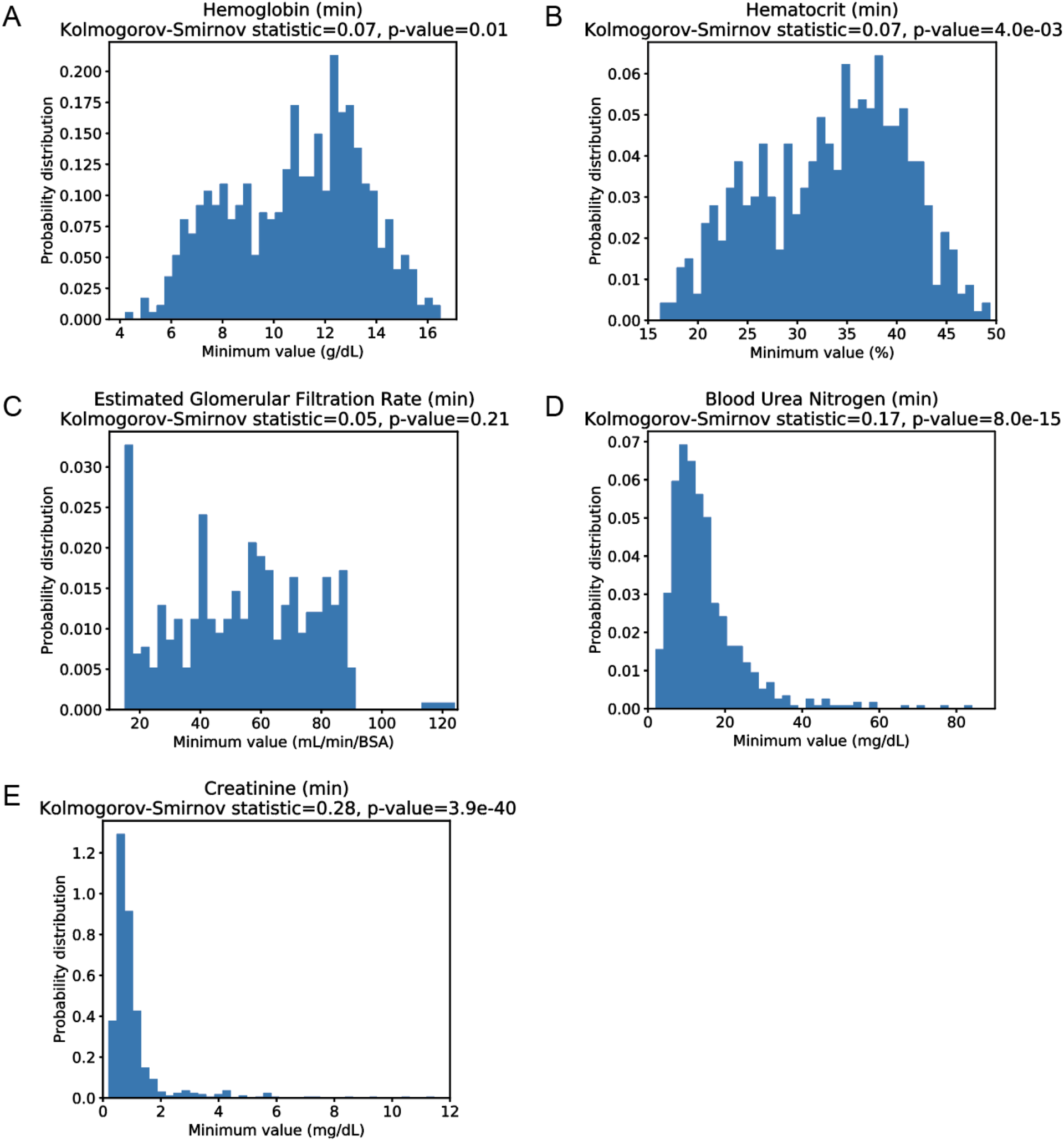
Distributions and normality tests of patient-wise minimum values for all considered lab tests. Lab tests evaluated include (A) hemoglobin, (B) hematocrit, (C) eGFR, (D) BUN, and (E) serum creatinine. Above each plot, the Kolmogorov-Smirnov statistic and associated p-value are shown; a p-value below 0.05 indicates that the data do not follow a normal distribution.

**Figure S10.**
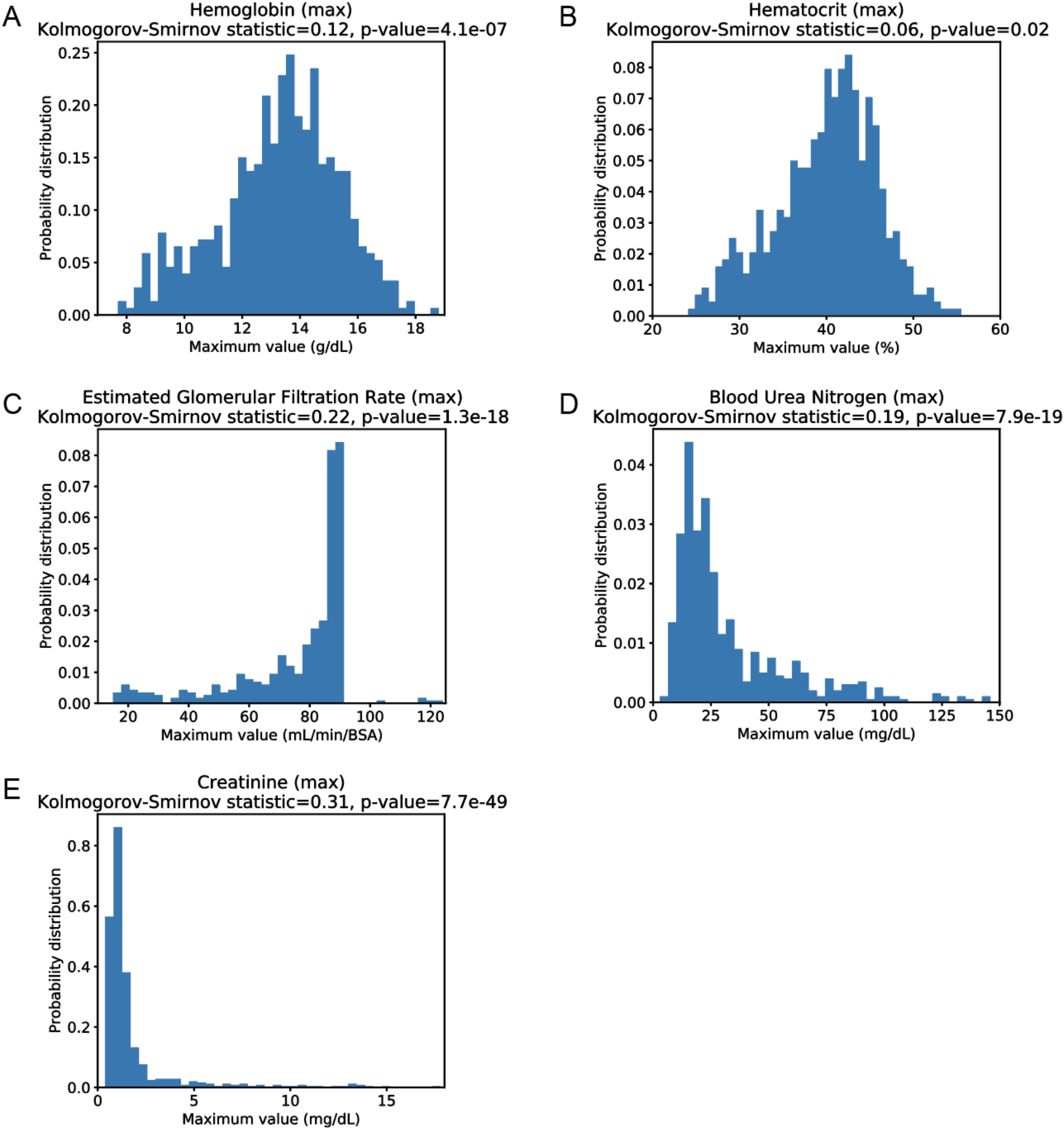
Distributions and normality tests of patient-wise maximum values for all considered lab tests. Lab tests evaluated include (A) hemoglobin, (B) hematocrit, (C) eGFR, (D) BUN, and (E) serum creatinine. Above each plot, the Kolmogorov-Smirnov statistic and associated p-value are shown; a p-value below 0.05 indicates that the data do not follow a normal distribution.

## References

Anand, P., Puranik, A., Aravamudan, M., Venkatakrishnan, A. J. and Soundararajan, V. (2020) ‘SARS-CoV-2 strategically mimics proteolytic activation of human ENaC’. eLife Sciences Publications Limited. doi: 10.7554/eLife.58603.

Bos, R., Rutten, L., van der Lubbe, J. E. M., Bakkers, M. J. G., Hardenberg, G., Wegmann, F., Zuijdgeest, D., de Wilde, A. H., Koornneef, A., Verwilligen, A., et al. (2020) ‘Ad26 vector-based COVID-19 vaccine encoding a prefusion-stabilized SARS-CoV-2 Spike immunogen induces potent humoral and cellular immune responses’, NPJ vaccines, 5, p. 91.

Carfì, A., Bernabei, R., Landi, F. and for the Gemelli Against COVID-19 Post-Acute Care Study Group (2020) ‘Persistent Symptoms in Patients After Acute COVID-19’, JAMA: the journal of the American Medical Association. American Medical Association, 324(6), pp. 603–605.

CDC (2020) COVID-19 and Your Health. Available at: https://www.cdc.gov/coronavirus/2019-ncov/need-extra-precautions/people-with-medical-conditions.html (Accessed: 19 January 2021).

Corbett, K. S., Flynn, B., Foulds, K. E., Francica, J. R., Boyoglu-Barnum, S., Werner, A. P., Flach, B., O’Connell, S., Bock, K. W., Minai, M., et al. (2020) ‘Evaluation of the mRNA-1273 Vaccine against SARS-CoV-2 in Nonhuman Primates’, The New England journal of medicine, 383(16), pp. 1544–1555.

Ellinghaus, D., Degenhardt, F., Bujanda, L., Buti, M., Albillos, A., Invernizzi, P., Fernández, J., Prati, D., Baselli, G., Asselta, R., et al. (2020) ‘Genomewide Association Study of Severe Covid-19 with Respiratory Failure’, The New England journal of medicine. N Engl J Med, 383(16). doi: 10.1056/NEJMoa2020283.

Folegatti, P. M., Ewer, K. J., Aley, P. K., Angus, B., Becker, S., Belij-Rammerstorfer, S., Bellamy, D., Bibi, S., Bittaye, M., Clutterbuck, E. A., et al. (2020) ‘Safety and immunogenicity of the ChAdOx1 nCoV-19 vaccine against SARS-CoV-2: a preliminary report of a phase 1/2, single-blind, randomised controlled trial’, The Lancet, 396(10249), pp. 467–478.

Jackson, L. A., Anderson, E. J., Rouphael, N. G., Roberts, P. C., Makhene, M., Coler, R. N., McCullough, M. P., Chappell, J. D., Denison, M. R., Stevens, L. J., et al. (2020) ‘An mRNA Vaccine against SARS-CoV-2 - Preliminary Report’, The New England journal of medicine. N Engl J Med, 383(20). doi: 10.1056/NEJMoa2022483.

Khwaja, A. (2012) ‘KDIGO Clinical Practice Guidelines for Acute Kidney Injury’, Nephron Clinical Practice. Karger Publishers, 120(4), pp. c179–c184.

Latz, C. A., DeCarlo, C., Boitano, L., Maximilian Png, C. Y., Patell, R., Conrad, M. F., Eagleton, M. and Dua, A. (2020) ‘Blood type and outcomes in patients with COVID-19’, Annals of hematology. Springer, 99(9), pp. 2113–2118.

Mercado, N. B., Zahn, R., Wegmann, F., Loos, C., Chandrashekar, A., Yu, J., Liu, J., Peter, L., McMahan, K., Tostanoski, L. H., et al. (2020) ‘Single-shot Ad26 vaccine protects against SARS-CoV-2 in rhesus macaques’, Nature. Nature Publishing Group, 586(7830), pp. 583–588.

Mohsenin, V. (2017) ‘Practical approach to detection and management of acute kidney injury in critically ill patient’, Journal of intensive care medicine. BioMed Central, 5(1), pp. 1–8.

Mulligan, M. J., Lyke, K. E., Kitchin, N., Absalon, J., Gurtman, A., Lockhart, S., Neuzil, K., Raabe, V., Bailey, R., Swanson, K. A., et al. (2020) ‘Phase I/II study of COVID-19 RNA vaccine BNT162b1 in adults’, Nature. Nature Publishing Group, 586(7830), pp. 589–593.

Pascarella, G., Strumia, A., Piliego, C., Bruno, F., Del Buono, R., Costa, F., Scarlata, S. and Agrò, F. E. (2020) ‘COVID-19 diagnosis and management: a comprehensive review’, Journal of internal medicine. J Intern Med, 288(2). doi: 10.1111/joim.13091.

Pawlowski, C., Venkatakrishnan, A. J., Ramudu, E., Kirkup, C., Puranik, A., Kayal, N., Berner, G., Anand, A., Barve, R., O’Horo, J. C., et al. (2020) ‘Pre-existing conditions are associated with COVID patients’ hospitalization, despite confirmed clearance of SARS-CoV-2 virus’, medRxiv. Cold Spring Harbor Laboratory Press, p. 2020. 10.28.20221655.

del Rio, C., Collins, L. F. and Malani, P. (2020) ‘Long-term Health Consequences of COVID-19’, JAMA: the journal of the American Medical Association. American Medical Association, 324(17), pp. 1723–1724.

Seabold, S. and Perktold, J. (2010) ‘Statsmodels: Econometric and Statistical Modeling with Python’, Proceedings of the 9th Python in Science Conference. doi: 10.25080/majora-92bf1922-011.

Singh, M., Bansal, V. and Feschotte, C. (2020) ‘A Single-Cell RNA Expression Map of Human Coronavirus Entry Factors’, Cell reports. Elsevier, 32(12), p. 108175.

Townsend, L., Dyer, A. H., Jones, K., Dunne, J., Mooney, A., Gaffney, F., O’Connor, L., Leavy, D., O’Brien, K., Dowds, J., et al. (2020) ‘Persistent fatigue following SARS-CoV-2 infection is common and independent of severity of initial infection’, PloS one. Public Library of Science, 15(11), p. e0240784.

Venkatakrishnan, A. J., Puranik, A., Anand, A., Zemmour, D., Yao, X., Wu, X., Chilaka, R., Murakowski, D. K., Standish, K., Raghunathan, B., et al. (2020) ‘Knowledge synthesis of 100 million biomedical documents augments the deep expression profiling of coronavirus receptors’. eLife Sciences Publications Limited. doi: 10.7554/eLife.58040.

Virtanen, P., Gommers, R., Oliphant, T. E., Haberland, M., Reddy, T., Cournapeau, D., Burovski, E., Peterson, P., Weckesser, W., Bright, J., et al. (2020) ‘Author Correction: SciPy 1.0: fundamental algorithms for scientific computing in Python’, Nature methods, 17(3), p. 352.

Wagner, T., Shweta, F. N. U., Murugadoss, K., Awasthi, S., Venkatakrishnan, A. J., Bade, S., Puranik, A., Kang, M., Pickering, B. W., O’Horo, J. C., et al. (2020) ‘Augmented curation of clinical notes from a massive EHR system reveals symptoms of impending COVID-19 diagnosis’. eLife Sciences Publications Limited. doi: 10.7554/eLife.58227.

Yelin, D., Wirtheim, E., Vetter, P., Kalil, A. C., Bruchfeld, J., Runold, M., Guaraldi, G., Mussini, C., Gudiol, C., Pujol, M., et al. (2020) ‘Long-term consequences of COVID-19: research needs’, The Lancet infectious diseases. Elsevier, 20(10), p. 1115.

Zhao, Y., Zhao, Z., Wang, Y., Zhou, Y., Ma, Y. and Zuo, W. (2020) ‘Single-Cell RNA Expression Profiling of ACE2, the Receptor of SARS-CoV-2’, American journal of respiratory and critical care medicine. American Thoracic Society, 202(5), p. 756.

Ziegler, C. G. K., Allon, S. J., Nyquist, S. K., Mbano, I. M., Miao, V. N., Tzouanas, C. N., Cao, Y., Yousif, A. S., Bals, J., Hauser, B. M., et al. (2020) ‘SARS-CoV-2 Receptor ACE2 Is an Interferon-Stimulated Gene in Human Airway Epithelial Cells and Is Detected in Specific Cell Subsets across Tissues’, Cell. Cell, 181(5). doi: 10.1016/j.cell.2020.04.035

